# The CSF-1R inhibitor Pexidartinib impacts dendritic cell differentiation through inhibition of FLT3 signaling and may antagonize the effect of durvalumab in patients with advanced cancer – results from a phase 1 study

**DOI:** 10.1101/2023.02.15.23285939

**Authors:** Aurélien Voissière, Carlos Gomez-Roca, Sylvie Chabaud, Céline Rodriguez, Axelle Nkodia, Justine Berthet, Laure Montane, Anne-Sophie Bidaux, Isabelle Treilleux, Lauriane Eberst, Catherine Terret, Iphigénie Korakis, Gwenaelle Garin, David Pérol, Jean-Pierre Delord, Christophe Caux, Bertrand Dubois, Christine Ménétrier-Caux, Nathalie Bendriss-Vermare, Philippe A. Cassier

## Abstract

Tumor-associated macrophages (TAM) are critical determinant of resistance to programmed death-1/programmed death-ligand 1 (PD-1/PD-L1) blockade. This phase I study (MEDIPLEX, NCT02777710) investigated the safety and efficacy of pexidartinib, a CSF-1R-directed tyrosine kinase inhibitor (TKI), and durvalumab (anti-PD-L1) in patients with advanced colorectal (CRC) and pancreatic (PDAC) carcinoma with the aim to enhance responses to PD-L1 blockade by eliminating CSF-1-dependent suppressive TAM. No unexpected toxicities were observed and 2% and 15% of patients achieved partial response and stable disease respectively. Increase of CSF-1 levels and decrease of CD14^low^CD16^high^ monocytes in peripheral blood mononuclear cells (PBMC) confirmed CSF-1R engagement. Treatment significantly decreased blood dendritic cell (DC) subsets and impaired IFN-λ/IL-29 production by type-1 conventional DC in *ex vivo* TLR3-stimulated PBMC. Pexidartinib also targets c-KIT and FLT3, both key growth factor receptors of DC development and maturation. In patients, FLT3-L levels increased with pexidartinib treatment. *In vitro*, pexidartinib impaired the FLT3-L but not GM-CSF-dependent generation of DC subsets from murine bone marrow progenitors. Our results demonstrate that pexidartinib, through the inhibition of FLT3 signaling, has deleterious effect on DC differentiation, which may explain the limited anti-tumor clinical activity observed in this study. This study suggests that inhibition of FLT3 should be taken into account when combining TKIs with immune checkpoint blockers.

**One-sentence summary:** Pexidartinib affects the development of dendritic cells

## INTRODUCTION

Colorectal cancer (CRC) and pancreatic ductal adenocarcinoma (PDAC) are the most common gastrointestinal cancers in Western countries and are both associated with significant morbidity and mortality. Although their outcome have improved over time, the cure rate and the long-term survival remain low for patients with metastatic disease: the 5-year survival is ?11% for CRC and ?6% for PDAC *(1–3)*.

Although immune checkpoint inhibitors (ICI), such as PD-1/PD-L1 blockers, have shown remarkable efficacy in several tumor types, their efficacy as single agent has been limited in both CRC and PDAC, with the remarkable exception of patients with high microsatellite instability (MSI-H) tumors *(4–6)*, despite significant PD-L1 expression in 15-40% of samples *(7, 8)*. This suggests that co-targeting alternative pathways of immune suppression may be required to increase efficacy. In line with this hypothesis, the tumor microenvironment (TME) has been shown to play a crucial role in PDAC and CRC progression, maintenance, and resistance to therapy. Several cell types present in the TME, such as fibroblasts and macrophages, have been shown to exert both tumor-promoting and tumor-constraining roles. Efficient targeting of these cell populations for therapy has, however proven difficult *(9, 10)*. Tumor-associated macrophages (TAMs) are associated with poor clinical outcome in most carcinomas, owing to their potential to promote angiogenesis, local invasion, tumor cell dissemination, extracellular matrix (ECM) remodeling, and immunosuppression *(11)*.

Colony-stimulating factor 1 receptor (CSF-1R) is a transmembrane receptor tyrosine kinase (RTK) that is widely expressed by monocytes, macrophages, and granulocytes *(12)*. CSF-1R acts as a cell-surface receptor for colony-stimulating factor 1 (CSF-1) and interleukin-34 (IL-34) *(13)*. In cancer, CSF-1R signaling facilitates survival, proliferation and differentiation of TAMs within the TME *(14)*. In addition, CSF-1 promotes an M2 immunosuppressive phenotype *(15)*. Furthermore, several types of solid tumors express high levels of CSF-1, which is correlated with poor survival *(14)*. Preclinical studies have shown that depleting TAMs using CSF-1R inhibitors can enhance the efficacy of PD-1/PD-L1 blockade in several cancer models, including PDAC *(16, 17)*. Several CSF-1 and CSF-1R inhibitors have entered clinical development *(18, 19)*, and comprise both monoclonal antibodies (mAbs) targeting CSF-1 or CSF-1R and small-molecule inhibitors or the CSF-1R tyrosine kinase (TKI) which in many cases also inhibit the related kinases c-KIT and FLT3 *(19)*.

PLX3397 (pexidartinib), an orally available CSF-1R, KIT, and FLT3 TKI, is safe and effective in reducing TAMs in selected solid tumor types *(20–22)*. We hypothesized that depleting TAMs in the TME with pexidartinib would restore T-cell mediated tumor clearance and improve response to durvalumab, an anti-PD-L1 mAb, in patients with advanced/metastatic CRC and PDAC.

In this study, despite a tolerable safety profile, the clinical activity of the combination was limited. In patients’ blood samples, we observed increased circulating CSF-1 as well as a decrease in circulating non-classical (nc)-monocytes CD14^low^CD16^high^, two well described biomarkers of CSF-1R blockade suggesting adequate pharmacological target engagement *(23)*. However, we also observed a rapid reduction of circulating dendritic cell (DC) subset numbers on treatment leading to decreased production of type III interferons (IFN) following *ex vivo* stimulation of PBMC with Toll Like Receptor (TLR) 3 ligand (poly(I:C) (pI:C)). This observation prompted us to investigate the role of FLT3 inhibition by pexidartinib, which also targets FLT3 (IC50 of 160 to 400 nmol/L), albeit with decreased potency compared to CSF-1R (IC50 of 13 nmol/L) *(24)*. We showed that pexidartinib increased plasma levels of FLT3-L in patients and prevented *in vitro* Flt3-L-dependent murine DC differentiation from hematopoietic progenitors at low concentration. Given the importance of DC in the regulation of anti-tumor immune responses, our findings may explain the limited clinical activity of the combination of pexidartinib and durvalumab observed in this study, and alerts on unwanted effects of combining TKIs with immune checkpoint blockers.

## RESULTS

### Patient Disposition and Baseline Characteristics

Between June 2016 and January 2019, 66 patients were screened and 47 were enrolled and treated at 2 comprehensive cancer centers in France: 19 in the dose escalation and 28 in the two expansion cohorts (CRC and PDAC) (Fig. 1A). Baseline characteristics of patients are summarized in table 1. Briefly, the majority of patients were male (n=27, 57%), with good performance status, the median age was 59 (range 43-75) years, and 24 patients had CRC and 23 had PDAC. Five patients with CRC (all enrolled in the dose-escalation part) had MSI-H tumors. The median number of prior line of therapy was 2 (range 1-7), and was higher for patients with CRC compared to those with PDAC, reflecting the higher number of available lines of therapy in this disease.

**Fig. 1:**
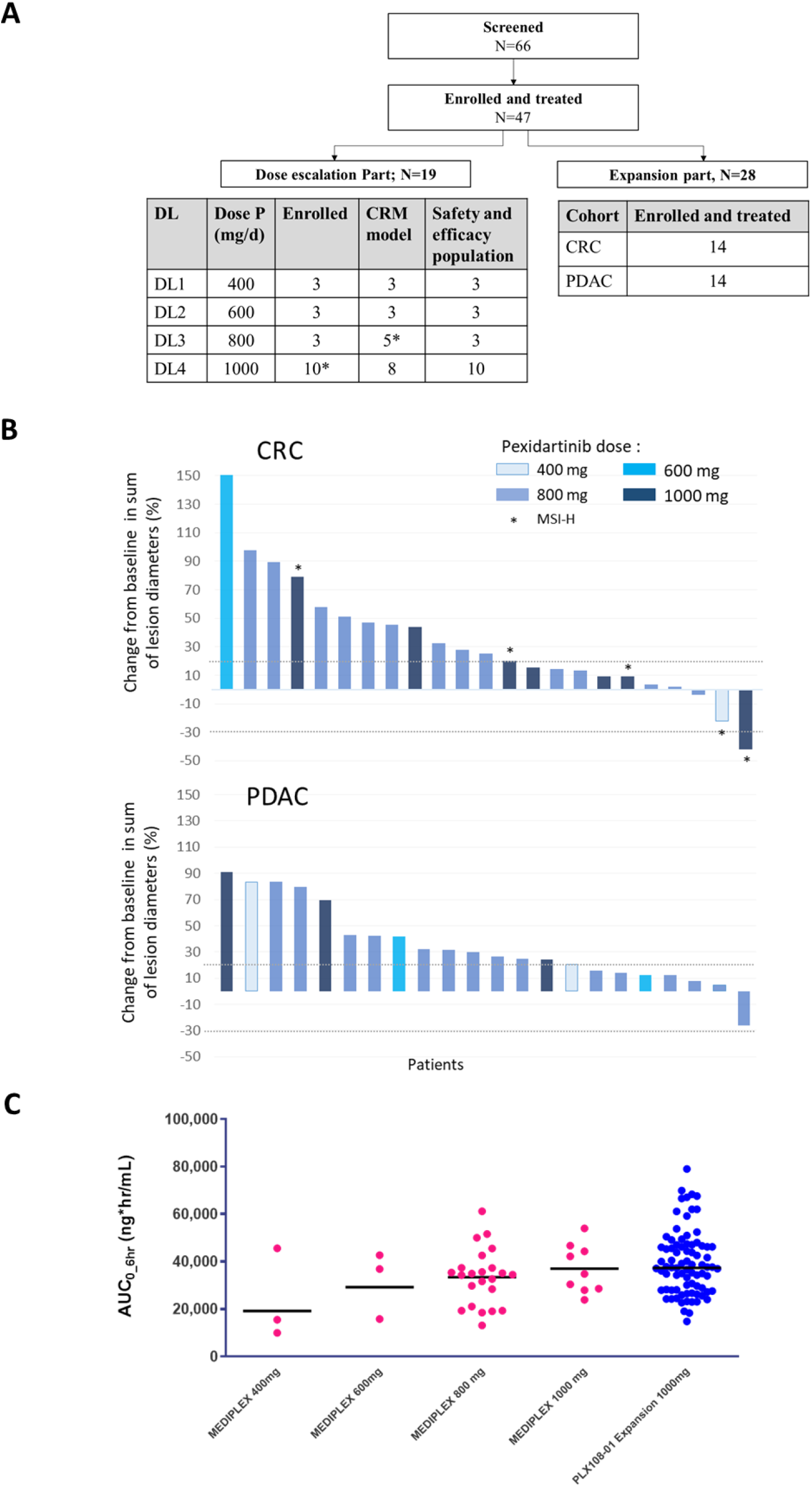
Consort, antitumor activity, and pharmacokinetic analysis of pexidartinib. **(A)** Study Consort diagram. *: Among 10 patients enrolled at 1000 mg/d DL, 2 patients have received during the DLT period a dose of pexidartinib equivalent/equal to 800 mg/d DL (i.e. one patient has received 93.5% of pexidartinib 800 mg/d DL, and the second patient has taken 800 mg/d during the Cycle 1 instead of 1000 mg/d) and were considered for the CRM model as part of DL3. Dose P, Dose Pexidartinib; DL, Dose level; CRC, Colorectal cancer; PDAC, Pancreatic ductal adenocarcinoma. **(B)** Waterfall plot. Best change from baseline in sum of RECIST target lesion diameters (%) for all enrolled patients (both parts) in the two cohorts (CRC and PDAC). Only 45 of 47 patients are shown; two patients (1 CRC and 1 PDAC) with no post-screening imaging were considered Non Evaluable for response. The dotted lines are positioned at +20% and −30%; these represent the thresholds for progressive disease and partial response per RECIST V1.1, respectively. **(C)** Steady-state systemic exposure (area under the curve from 0 to 6 hours [AUC0–6hr]) by dose level and in comparison with PLX108-01 study (Pexidartinib monotherapy; NCT01004861 *(20)*). At 800 mg/d, the patients in the escalation and expansion part are represented. Dots represent individual patients, whereas bars indicate geometric mean values.

**Table 1.**
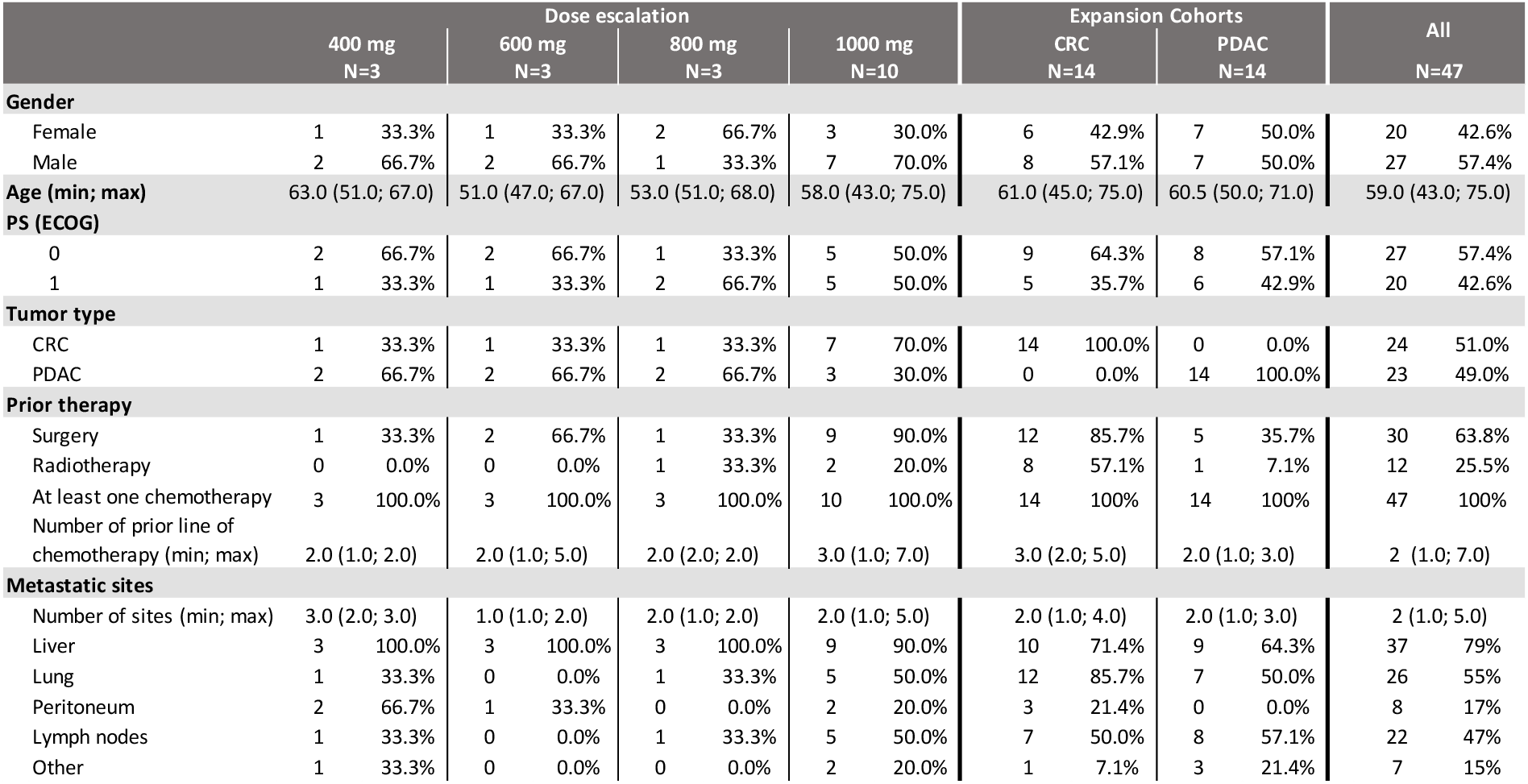
Patient’s characteristics at baseline. Data are n (%) or median (min; max). ECOG, Eastern Cooperative Oncology Group; PS, performance status; CRC, Colorectal cancer; PDAC, Pancreatic ductal adenocarcinoma.

### Safety, Dose-limiting toxicities, and recommended phase 2 dose

The safety profile of the combination was overall consistent with the expected adverse event (AE) profile of each drug, and no new safety signal was observed. In the dose escalation part, 18 patients (95%) experienced at least one treatment-related AEs (TRAEs) and 7 patients (37%) had at least one TRAE of Grade ≥ 3 according to NCI-CTCAE V4.03. In the expansion part, 27 patients (96%) experienced at least one TRAE and 15 patients (54%) had at least one TRAE of Grade ≥ 3. The most common all grade clinical TRAEs were fatigue, peripheral edema, rash, diarrhea, nausea, anorexia, vomiting, hair color changes and pruritus, all observed in more than 25% of treated patients, and were in most cases Grade 1-2 (table S1). The most common laboratory TRAEs were liver function test (LFT) abnormalities (increases in aspartate aminotransferase (AST), alanine aminotransferase (ALT), Alkaline phosphatase (ALP) and bilirubin), followed by decreased blood cell counts (anemia, absolute neutrophil and lymphocyte counts) (table S1). Increases in AST and ALT were in general rapidly reversible (<7 days) with interruption of pexidartinib and in most cases without the use of corticosteroids. Pexidartinib was escalated from 400 mg per day (400 mg/d) (200 mg [*bis in die* (BID)]) to 1000 mg/d (400 mg in the morning and 600 mg in the evening). No dose-limiting toxicities (DLT) were observed up to dose level (DL) 3 (800 mg/d). A total of 10 patients were enrolled at the 1000 mg/d DL (DL4). Two of these patients who received less than 80% of the planned pexidartinib dose during Cycle 1 for reasons other than toxicity were considered non-evaluable for DLT at this DL (1000 mg/d), but were included in the safety analysis (including in the CRM modeling) as being treated at 800 mg/d DL (Fig. 1A). Two other patients experienced DLT at 1000 mg/d DL, in both cases Grade 3 AST and ALT elevation, including one case with concomitant >2ULN bilirubin increase, deemed related to pexidartinib. At the end of 1000 mg/d DL (N= 8/19 patients treated and evaluable in the dose-escalation part), the CRM simulations indicated that the probability that the next 6 patients will be treated at the same DL (e.g. 1000 mg/d DL) was 100%, which was a pre-specified rule to stop dose-escalation. Due to the higher incidence of Grade ≥ 3 TRAEs (mainly LFT abnormalities – table S1) and toxicity-related dose modifications (table S2) at 1000 mg/d DL, the recommended dose for expansion cohorts (RP2D) was pexidartinib 800 mg/d (400 mg BID) with durvalumab (1500 mg/4 weeks, IV).

### Treatment exposure, dose modifications, and interruptions

The median durations of treatment with pexidartinib were 1.8 (range 0.8-18.4) months and 1.7 (range 0.6-3.7) months in the dose escalation and expansion cohorts respectively, while durations of treatment with durvalumab were 1.8 (range 0.9-33.1) and 1.8 (range 0.9-8.9) months respectively. Overall, 34 patients (72%) had at least one dose modification of pexidartinib, due to AEs for 61% of them. In most cases treatment interruption and dose-modification were done to manage AST/ALT and/or bilirubin increase, nausea/vomiting, maculopapular rash, and neutropenia (table S2). All patients but one permanently discontinued both study drugs. One patient with MSI-H CRC continued treatment with single agent durvalumab for 33 months. The most common reason for permanent discontinuation of study treatments was disease progression (94%). However, 7 patients (15%) permanently discontinued pexidartinib due to AE: 7 patients for Grade 3 AST/ALT increase including 3 cases with concomitant Grade 2 bilirubin increase, one patient for Grade 4 neutropenia and one patient for Grade 3 maculopapular rash. The causes of permanent treatment discontinuations are summarized in table S2.

### Clinical activity

The best overall response per RECIST v1.1 was PR in one patient with MSI-H CRC, while 7 patients had SD and 39 had PD. Among the 7 patients qualified as SD, two patients had tumor shrinkage beyond 10%: one patient with MSI-H CRC had prolonged SD (−22%) lasting more than 15 months; one patient with PDAC had a 26% tumor shrinkage that lasted 3.6 months (Fig. 1B). The median PFS in the expansion cohorts (n=28) was 1.8 months and was similar among patients with CRC and PDAC, while, the OS was longer for CRC patients (Fig. S1).

### Pharmacokinetics

Plasma samples from all 47 patients were available for pharmacokinetic analysis of pexidartinib. Despite the expected variability, AUC0-6 (area under the concentration-time curve from time zero to 6 hours) increased in a dose proportional manner, with values that were comparable to those observed in the single agent phase 1 study (PLX108-01 study; NCT01004861) *(20)*. At steady state (C1D15), at the RP2D (800 mg/day), the AUC0-6 was 31300 ng*hr/mL, which did not differ significantly from the values observed at 1000 mg/day (35800 ng*hr/mL for the AUC0-6) (Fig. 1C). The mean plasma half-life in the dose expansion was 7.23 h, consistent with previous data obtained in the single agent phase 1 study *(20)*. Overall, this suggests that durvalumab had no impact on pexidartinib exposure.

### Pharmacodynamic biomarkers

Target engagement of pexidartinib was assessed by investigating circulating levels of CSF-1 and of nc-monocytes (CD14^low^CD16^high^) which rely on CSF-1R signaling for their survival *(25)*. CSF-1 levels in plasma increased after pexidartinib treatment in all 47 patients, in a dose- and time-dependent manner up to C1D15, as previously reported *(26)*. At 800 mg/day, a 5-fold increase in the mean CSF-1 levels was observed at C1D15 as compared to C1D1 (Fig. 2A) without any difference between CRC or PDAC patients. Elevated CSF-1 levels were maintained beyond C1D15 in patients who continued to receive pexidartinib, but rapidly returned to baseline levels in patients who interrupted or permanently discontinued pexidartinib (Fig. S2). At C2D1, plasma levels of pexidartinib and CSF-1 were correlated (r^2^ = 0.53, p = 0.0006) (Fig. 2B). Furthermore, pexidartinib treatment induced a 10-fold reduction in circulating CD14^low^CD16^high^ nc-monocytes between baseline and C1D15 (Fig. 2C), as previously reported *(26)*. In contrast, classical monocytes (CD14^high^CD16^low^) and inflammatory monocytes (CD14^high^CD16^high^) were not affected (Fig. 2C). The upregulation of circulating CSF-1 concentrations and the reduction of CSF-1-dependent cell numbers are consistent with CSF-1R blockade by pexidartinib. We also observed a transient increase of IFN-γ in plasma at C1D15 (Fig. 2D left) reflecting PD-1/PD-L1 signaling blockade by durvalumab. Of note, the dose of pexidartinib did not significantly alter durvalumab-triggered IFN-γ elevation in patients’ plasma at C1D15 (Fig. 2D right).

**Fig. 2:**
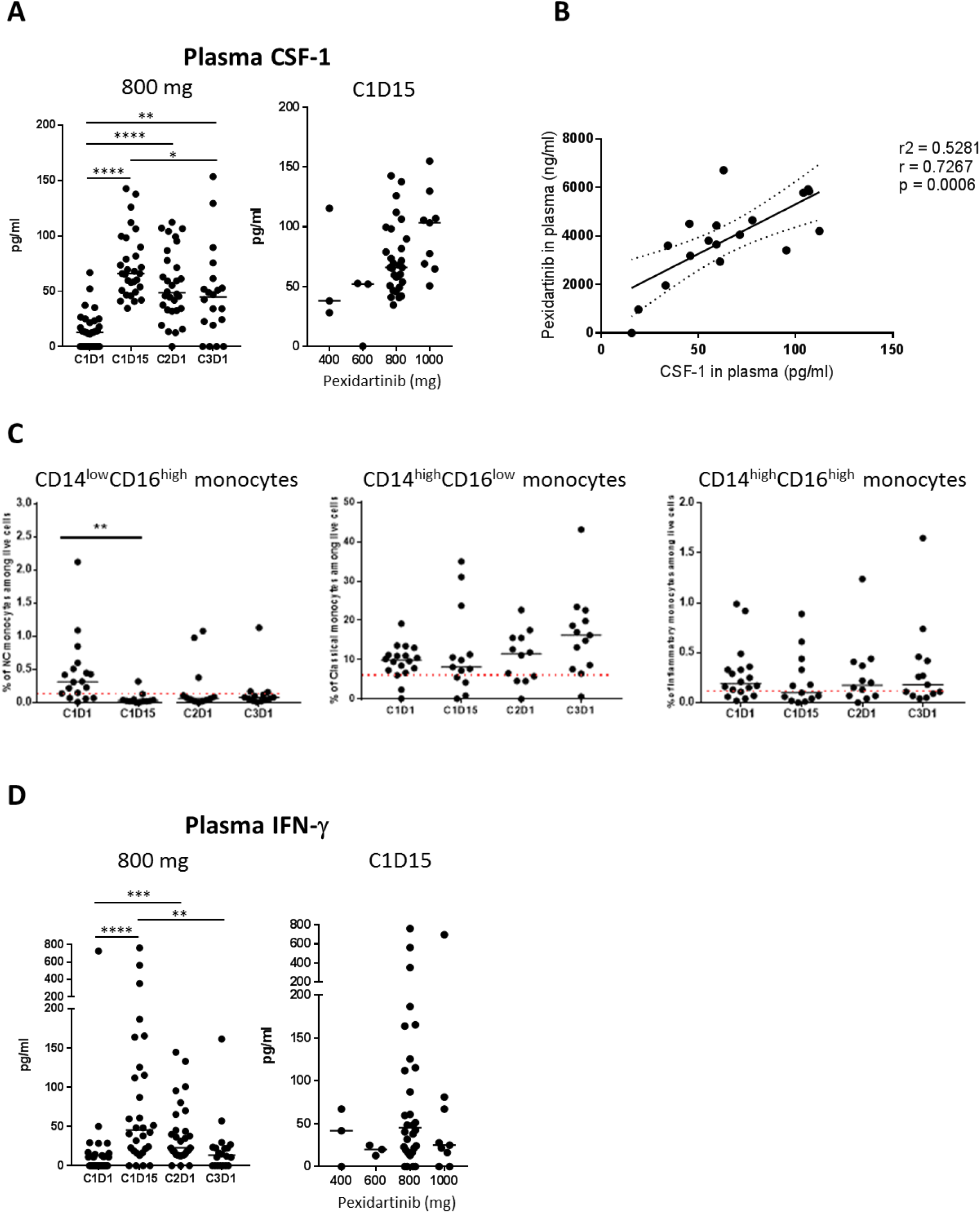
Treatment with pexidartinib increased the plasma CSF-1 levels and decreased the proportions of circulating nc-monocytes. (**A**) CSF-1 levels were quantified by multiplex ECLIA in plasma of patients from the expansion cohort (**left panel**) and the dose escalation at C1D15 (**right panel**). Dots represent individual patients, whereas bars indicate median values. (**B**) Correlation between pharmacokinetic of pexidartinib and plasmatic CSF-1 levels at C2D1 (linear regression test). (**C**) Phenotypic analyses of frozen patients’ PBMCs by FC to assess the modulation of proportions of CD14^high^CD16^low^, CD14^high^CD16^high^, and CD14^low^CD16^high^ monocyte subsets among PBMC. Dots represent individual patients, whereas bars indicate median values. Red dotted lines represent healthy donors’ median on each graph. (**D**) IFN-γ levels were quantified by multiplex ECLIA in plasma of patients from the expansion cohort (**left panel**) and the dose escalation at C1D15 (**right panel**). Dots represent individual patients, whereas bars indicate median values. Data were analyzed using a Kruskal-Wallis test followed by Dunn’s correction. *: *P* < 0.05; **: *P* < 0.01; ***: *P* < 0.001; and ****: *P* < 0.0001.

Only 10 of 28 patients (36%) enrolled in the expansion phase had paired pre- and on-treatment tumor biopsies assessable to investigate changes in immune infiltrate *in situ*. In the remaining 18 patients, the amount of tumor in the on-treatment biopsy was insufficient for 11 patients (39%) or not done for 7 patients due to disease progression (25%). Among the 10 patients with paired tumor samples, we were unable to detect robust changes in TAMs (CD68/CD163), polymorphonuclear cells (PMN), myeloperoxydase (MPO)), and T cells (CD3). Nevertheless, one PDAC patient (01-032) who had significant, albeit transient, tumor shrinkage (Fig. 3A, 3B) had signs of pre-existing anti-tumor immunity as demonstrated by the presence of high infiltrate in PMNs (Fig. 3C) and TAMs (Fig. 3E), tertiary lymphoid structures based on T and B cell infiltrate organization (Fig. 3G), and proliferating (Ki67^+^) T cells (Fig. 3I) on the pre-treatment biopsy. In this particular case, treatment led to decrease in PMN and TAM infiltration (Fig. 3D, 3F) and to a lesser extent in T-cell infiltration with reduced proliferation (Fig. 3H, 3J).

**Fig. 3:**
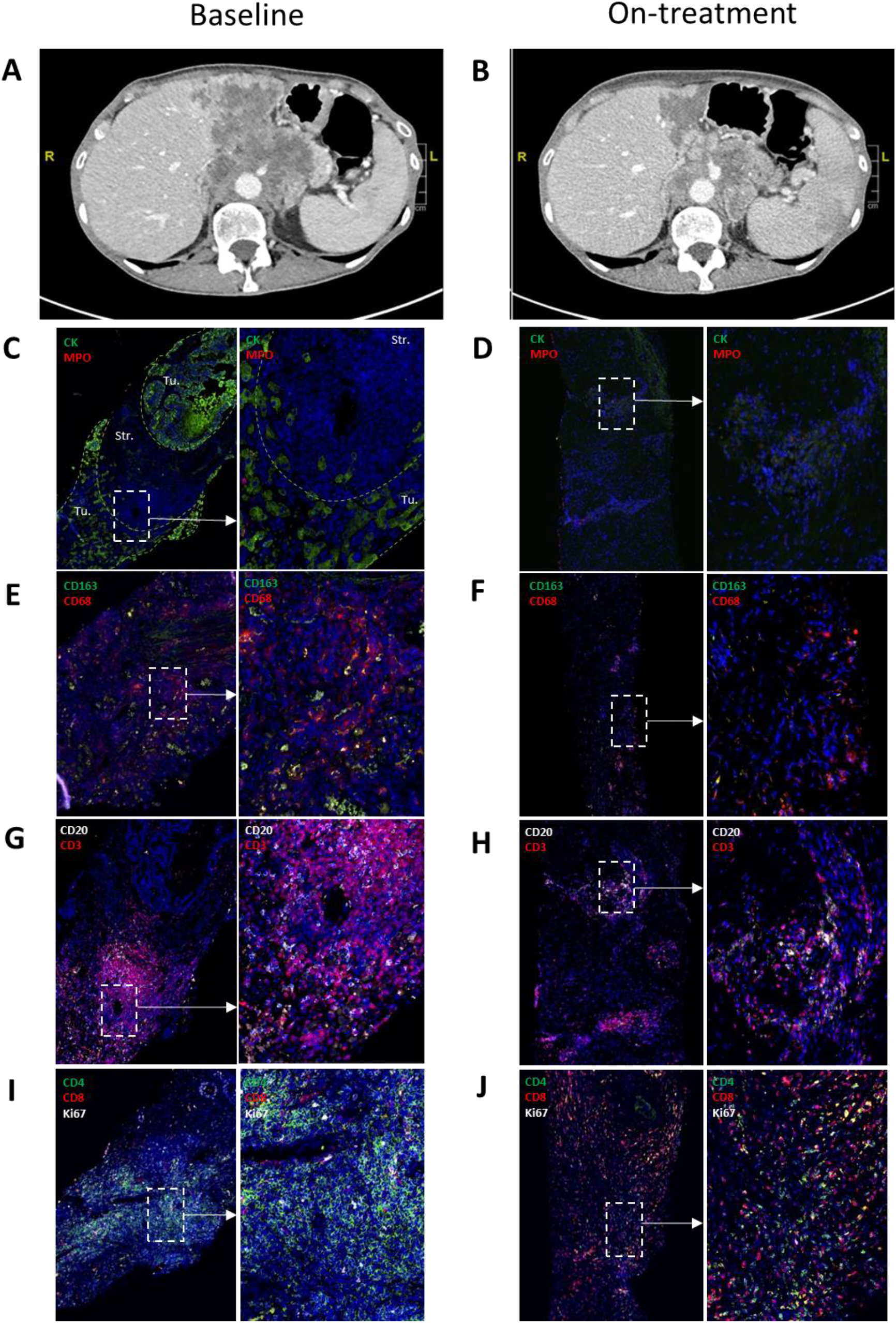
Evidence of pre-existing lymphocytic immune infiltrate in a patient with stable disease. For the PDAC patient (#01-032), tumor scan demonstrating the tumor shrinkage comparing (**A**) inclusion and (**B**) on treatment. (**C-J**) Representative illustrations of (**C, D**) CK-MPO, (**E, F**) CD68-CD163 (**G, H**) CD3-CD20, and (**I, J**) CD4-CD8-Ki67 serial multi-IF stainings of FFPE tumor tissue biopsies collected at baseline (**C, E, G, I**) and at C2D1 (**D, F, H, J**). Nuclei were stained with DAPI (blue). Dotted lines in A delineate the tumor and stromal areas based on CK staining.

### Pexidartinib reduces the frequency of all circulating DC subsets in patients

Aside from IFN-γ increase (Fig. 2D), we did not detect any other change in cytokines associated with T cells (IL-2, IL-13, IL-17A) or innate immune responses (GM-CSF, IFN-α, IL-1β, IL-12p70, and TNF-α) in patients’ plasma at baseline or on treatment. Furthermore, we assessed the proportions of immune cell subpopulations in blood as well as their functionality following *ex vivo* short-term stimulation by multiparametric flow cytometry (FC) analyses for patients enrolled in the expansion phase, as previously described *(27)*. TLR7/8 (R848) stimulation triggered the production of inflammatory cytokines (IL-1β, IL-6, TNF-α), that was not modulated by the treatment over time (Fig. S3A). Furthermore, no significant changes in the frequency of T cells and CD4/CD8 ratio (Fig. S3B) and in T cell functionality (IFN-γ and IL-2 production) after short term PMA-ionomycin reactivation (Fig. S3C) were noted on treatment compared to baseline. A very transient decrease was observed for B cells at C1D15 (Fig. S3B). In contrast, we observed a significant decrease of all circulating DC subsets, namely BDCA2^+^ plasmacytoid DC (pDC), type 1 conventional BDCA3^+^ dendritic cells (cDC1), and type 2 conventional BDCA1^+^DC (cDC2) at C1D15 compared to C1D1 (Fig. 4A). This was accompanied by a drop in IL-29/IFN-λ levels, known to be specifically produced by cDC1 *(28)*, in the supernatants of pI:C-stimulated PBMC at C1D15, while the modulation of IFN-α which is mainly secreted by pDC in response to R848 *(27)* was less obvious due to the alteration of basal pDC function in these patients (Fig. 4B).

**Fig. 4:**
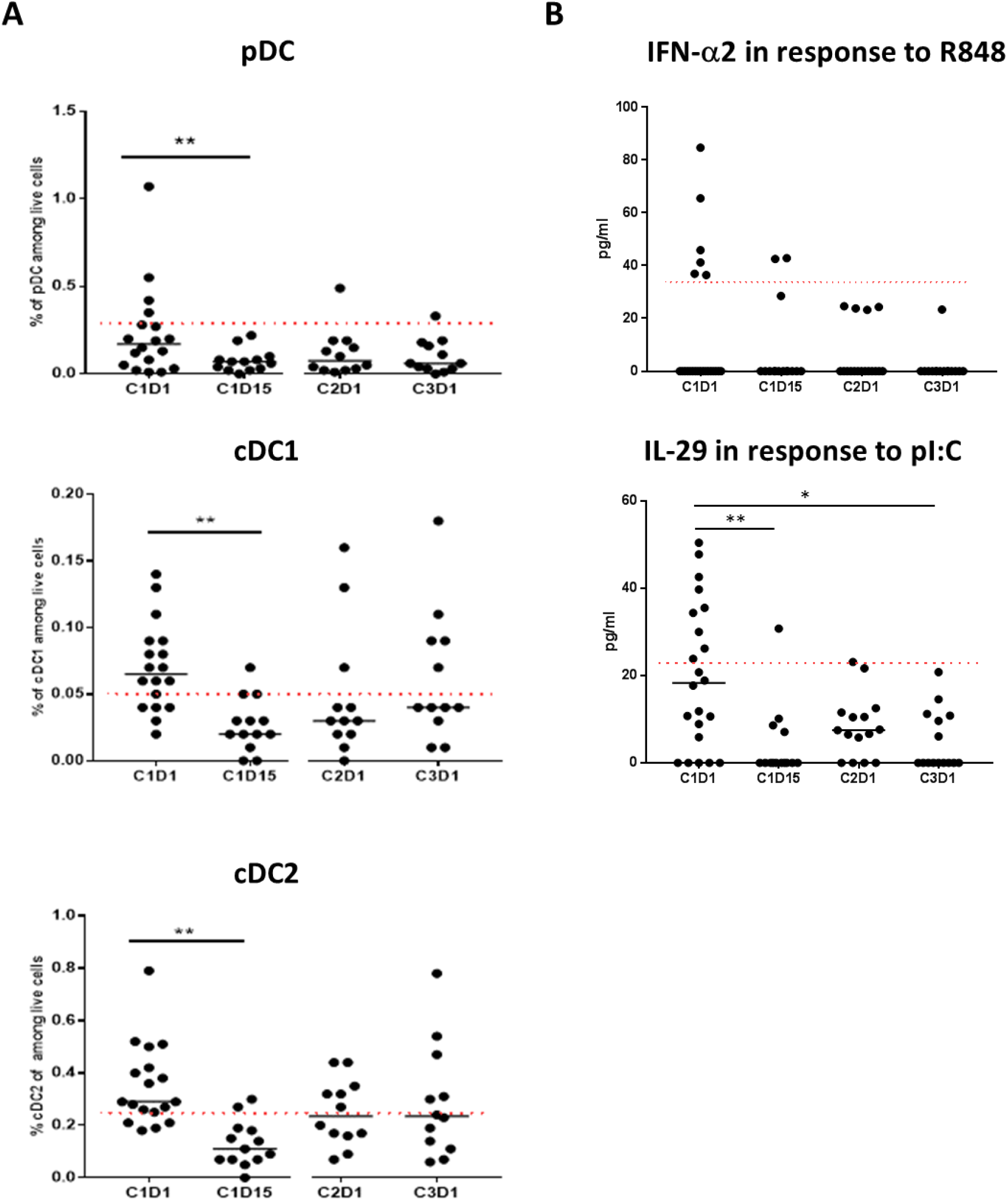
Treatment with pexidartinib and durvalumab combination reduced the proportion of circulating DC subsets. (**A**) Phenotypic analyses of frozen patients’ PBMCs by FC (panel 1, **table S3**) to assess the proportions of pDC, cDC1, and cDC2 among live cells as previously described *(27)*. Dots represent individual patients, whereas bars indicate median values. Red dotted lines represent healthy donors’ median on each graph. (**B**) Quantification by multiplex ECLIA of IFNα-2a and IL-29 in supernatants of patients’ PBMCs after 6 hours stimulation with TLR-L (respectively R848 and pI:C). Dots represent individual patients, whereas bars indicate median values. Data were analyzed using a Kruskal-Wallis test followed by Dunn’s correction. Red dotted lines represent healthy donors’ median on each graph. *: *P* < 0.05; **: *P* < 0.01.

### Pexidartinib increases circulating levels of FLT3-L

Pexidartinib does not only inhibit CSF-1R (IC50=13nM) but also other related RTK such as c-KIT (SCF receptor) (IC50=27nM) and FLT3 (IC50=160nM) *(20, 29)*, which are both key growth factor receptors for DC differentiation, maturation, and viability (for review *(30)*). Therefore, we assessed changes in the plasma levels of their cognate ligands, SCF and FLT3-L during treatment. Whereas SCF levels were not modified by treatment (Fig. 5A), we observed a sustained increase (by 2-fold) in circulating FLT3-L levels in the expansion cohort (Fig. 5B), pointing to a systemic impact of pexidartinib on FLT3 signaling. Moreover, this increase was dependent on the dose of pexidartinib (Fig. 5B), but, unlike what was observed for CSF-1 (Fig. 2A), plasma levels of FLT3-L did not correlate to those of pexidartinib (r^2^=0.053, p=0.41) (Fig. 5C). Taken together these data suggest that, despite a higher IC50 reflecting lower activity of pexidartinib on FLT3, pexidartinib modulates FLT3 pathway at therapeutically relevant doses in human.

**Fig. 5:**
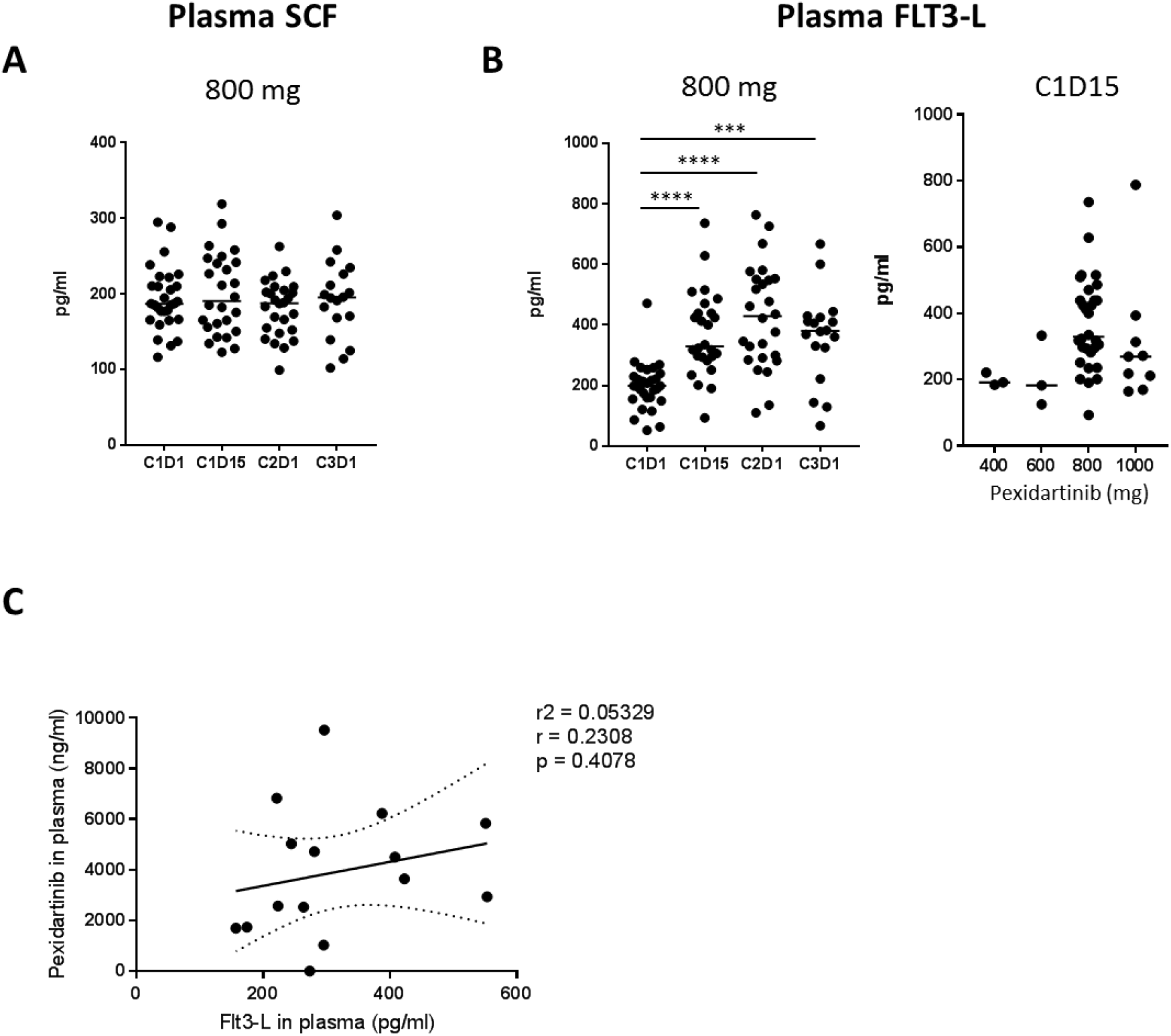
Treatment with pexidartinib increased FLT3-L but not SCF plasma levels. (**A**) SCF and (**B**) FLT3-L levels were quantified by multiplex ECLIA in plasma of patients from the expansion cohort (**A** and **B, left panel**) and the dose escalation at C1D15 (**B, right panel**). Dots represent individual patients, whereas bars indicate median values. Data were analyzed using a Kruskal-Wallis test followed by Dunn’s correction. (**C**) Correlation between pharmacokinetic of pexidartinib and plasma FLT3-L levels at C2D1 (linear regression test).

### Inhibition of FLT3 by pexidartinib impairs murine DC differentiation in bone marrow cultures

To investigate the impact of pexidartinib on DC differentiation, we used well-established *in vitro* culture systems set up with murine bone marrow (BM) progenitors and supported by Flt3-L *(31)* or GM-CSF *(32)*. BM cells were cultured in the presence of Flt3-L or GM-CSF for 7 days with increasing doses of pexidartinib followed by FC analysis. Total DC were identified as CD11c^+^ cells among cultured cells and then scored for XCR1, SIRPα, CD11b, and SiglecH expression to identify cDC1 (XCR1^+^ SIRPα^neg^), cDC2 (XCR1^neg^ SIRPα^+^), and pDC (SiglecH^+^, Bst2^+^ CD11b^neg^) (Fig. 6A). In this model, pexidartinib inhibited Flt3-L-driven differentiation of total CD11c^+^ DC in a dose-dependent manner, whether using recombinant Flt3-L (rFlt3-L) or B16-Flt3-L culture supernatant (sFlt3-L) (Fig. 6B). In contrast, pexidartinib had no impact on GM-CSF-driven DC generation (Fig. 6B). Furthermore, when added at the beginning of the culture (day 0), pexidartinib (1μM) reduced the percentages and numbers of each DC subset that were differentiated with rFlt3-L but had a weaker impact when added in culture at day 2 or 4 on cells already engaged in DC differentiation process (Fig. 6B). Thus, the negative impact of pexidartinib on Flt3-L-driven DC generation declined over time. Finally, pexidartinib at 10μM (a clinically relevant concentration) strongly reduced cDC1 and cDC2 viability, while the viability of pDC was inhibited only at the highest dose of 40μM (Fig. 6C). In human, pexidartinib did not significantly alter the survival of blood DC subsets, although there was a trend for pDC at 40μM as for mouse pDC (Fig. S4). Taken together, these results show that, at therapeutically relevant doses, pexidartinib acts mainly on DC differentiation from progenitors rather than on survival of fully differentiated DC *in vitro*.

**Fig. 6:**
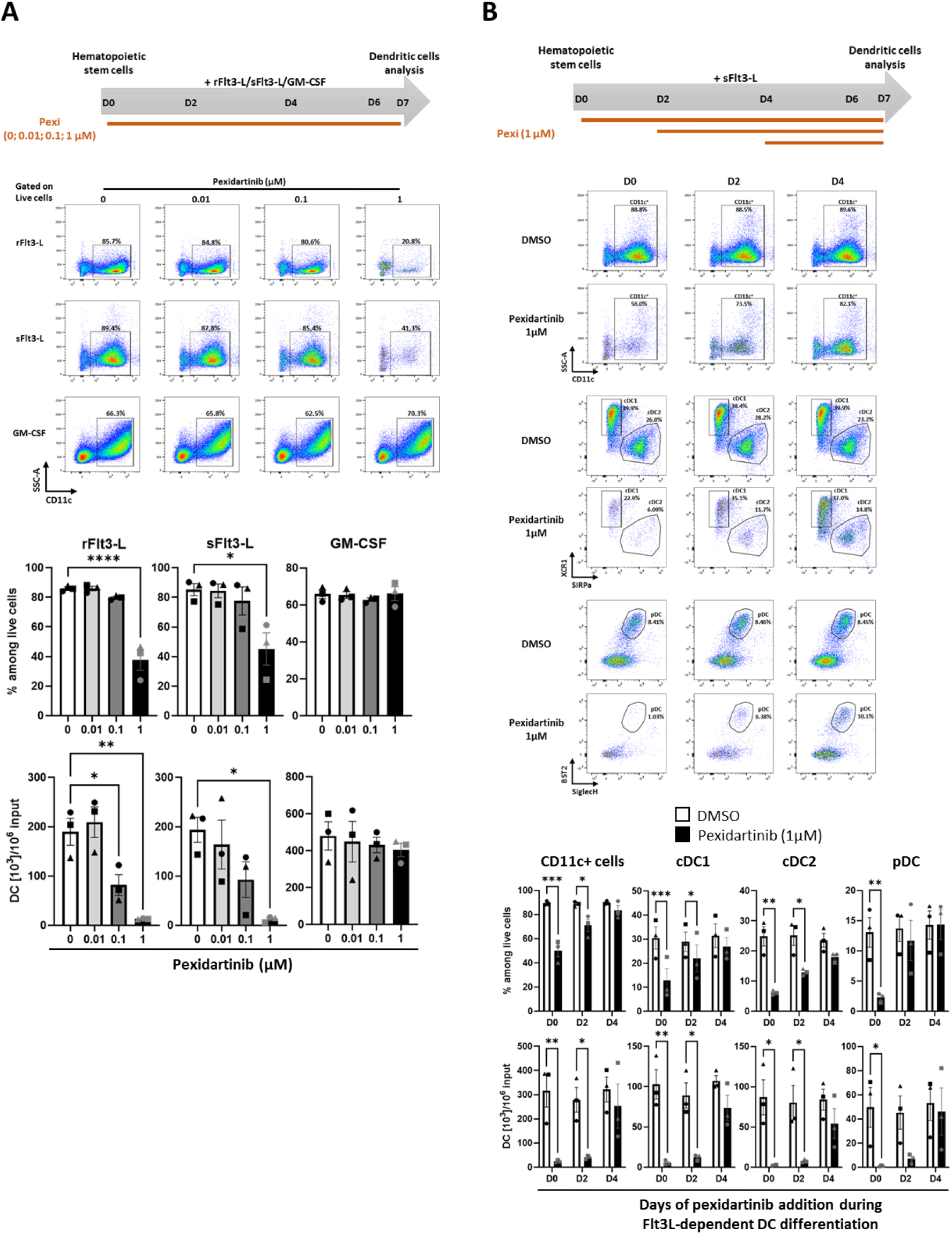
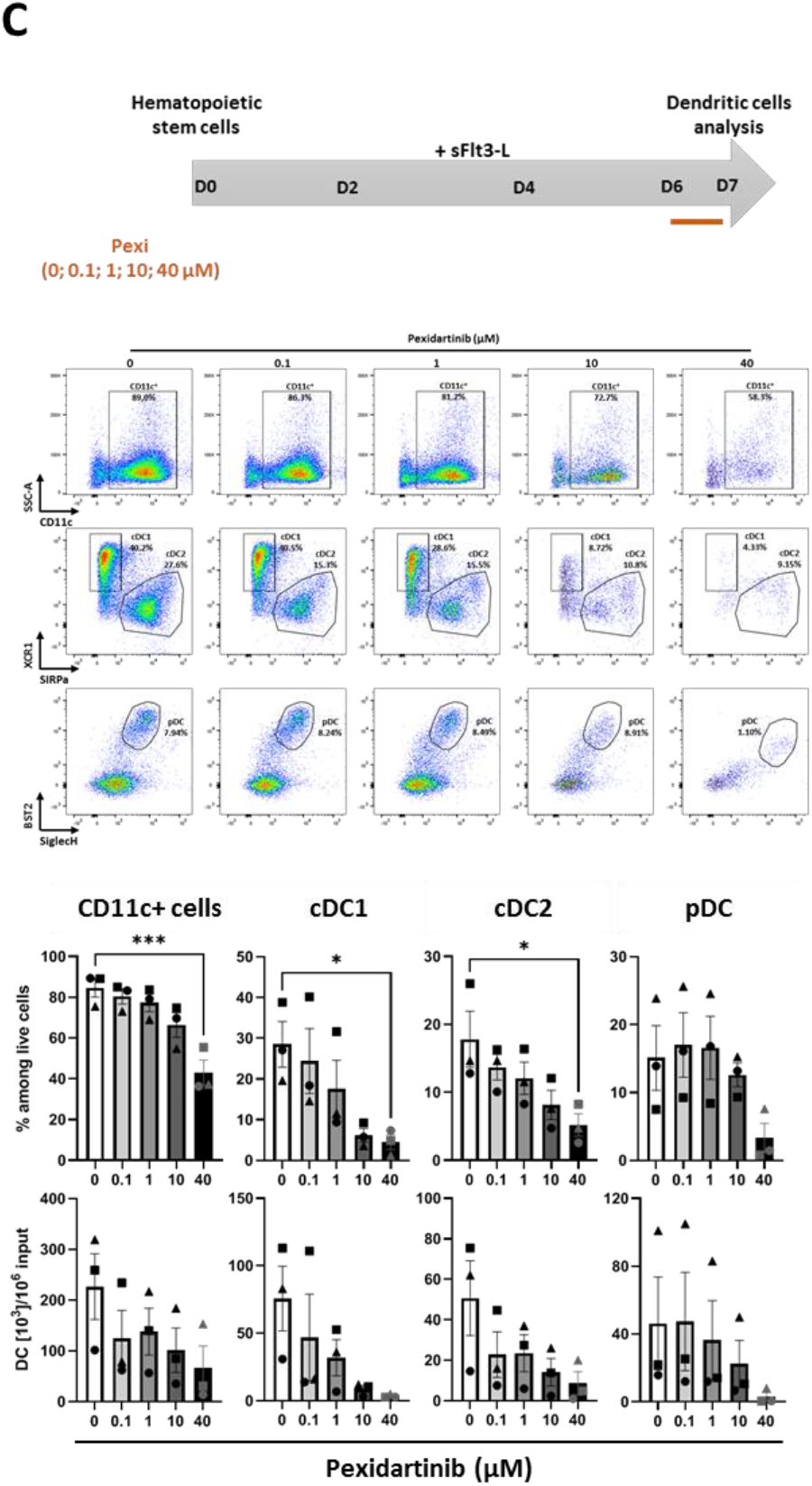
Treatment with pexidartinib altered the differentiation and survival of murine BM-DC. **(A)** Dose effect of pexidartinib on the differentiation of BM-DC. BM cells were exposed to cytokines (rFlt3-L, sFlt3-L or GM-CSF) and increasing doses of pexidartinib (0, 0.01, 0.1, 1 μM) for 7 days. Total DCs were identified as CD11c^+^ cells by FC at D7. Representative staining plots of differentiated CD11c^+^ DCs are shown. Histogram bars represent the fractions of CD11c^+^ DCs among live cells and the absolute numbers of CD11c^+^ DCs per 10^6^ initial undifferentiated BM cells. **(B)** Kinetic impact of pexidartinib on the differentiation of BM-DC. BM cells were exposed to sFlt3-L from D0 to D7 and pexidartinib (1 μM) or DMSO used as control was added at D0, D2 or D4 until D7. DCs were identified by FC as CD11c^+^ cells, cDC1 and cDC2 as XCR1^+^ SIRPα^neg^ and XCR1^neg^ SIRPα^+^ respectively among live CD11c^+^ MHC-II^+^ cells, and pDCs as BST2^+^ SiglecH^+^ among live CD11c^+^CD11b^neg^ cells at D7. Representative staining plots of differentiated total CD11c^+^ DCs, cDC1/cDC2, and pDCs are shown. Histogram bars represent the fractions of each population among live cells and the absolute numbers of each population per 10^6^ initial undifferentiated cells. **(C)** Dose-dependent impact of pexidartinib on the survival of fully Flt3L-differentiated BM-DC. BM-DC were differentiated from BM cells in the presence of sFlt3-L for 6 days. At D6 increasing doses of pexidartinib (0, 0.1, 1, 10 or 40 μM) were added for additional 18h of culture. DCs were identified as in panel (**B**). Representative staining plots of differentiated total CD11c^+^ DCs, cDC1/cDC2, and pDCs are shown. Histogram bars represent the fractions of each population among live cells and the absolute numbers of each population per 10^6^ initial undifferentiated cells. For each panel, data represent mean ± SEM of 3 independent cultures (each symbol represents the mean of duplicate in one experiment). Data were analyzed using a one-way ANOVA followed by Dunnett’s multiple comparisons test (**A, C**) and a 2-way ANOVA followed by Bonferroni’s multiple comparisons test (**B**). *: *P* < 0.05; **: *P* < 0.01; ***: *P* < 0.001; and ****: *P* < 0.0001.

## DISCUSSION

Eliminating tumor-promoting TAMs through the targeting of CSF-1/CSF-1R axis was suggested as a potential strategy for reversing tumor-induced immune suppression and enhancing the response to PD-(L)1 blockade in patients with cancer. In the present study, we investigated the safety and efficacy of pexidartinib, a TKI targeting CSF-1R, in combination with durvalumab, a PD-L1 blocker, in patients with advanced CRC or PDAC. The combination proved to have an acceptable toxicity profile but limited clinical activity. In addition, we showed that treatment with pexidartinib reduces circulating DC subset frequencies in patients, likely through inhibition of FLT3 signaling. *In vitro* experiments confirmed that pexidartinib inhibited *in vitro* differentiation of murine BM-DC. Given the importance of DC in anti-tumor immune response *(33, 34)* and in therapeutic response to anti-PD(L)1 *(35)*, this novel and key finding may explain the limited anti-tumor clinical activity observed in this study.

Similar to other studies with CSF-1R inhibitors, AST/ALT increase and edema were among the most common TRAEs. Transaminase elevation, together with increases in creatinine kinase and lactate dehydrogenase, result from decreased clearance due to the depletion of Kupffer cells, which are resident macrophages present in the liver sinusoids and depend on CSF-1R signaling *(36)*. The edema has been attributed to changes in ECM composition *(37, 38)*. Consistent with the previously described safety profile of pexidartinib, hair color changes and low grade gastrointestinal AE were also frequent *(39)*. Liver enzymes abnormalities of Grade 3 occurred in 20% of patients overall, but were all rapidly reversible following pexidartinib dose-interruption without the use of corticosteroids, suggesting they are pexidartinib-mediated (as opposed to immune-mediated hepatitis which is more commonly attributed to PD-(L)1 blockers). Here again, this is consistent with the known safety profile of pexidartinib, although the rate of grade 3 LFT abnormalities here was slightly higher than previously reported, albeit in a different patient population *(39)*. The rate of cutaneous AEs (rash and pruritus), including Grade 3 rashes in almost 10 % of patients, was higher than that reported for either pexidartinib or durvalumab as single agents *(39, 40)* suggesting a potential synergy between the two drugs. This is however consistent with other studies investigating CSF-1R inhibitors combined with PD-(L)1 blockers *(37, 41, 42)*.

We confirmed pharmacodynamics effects of pexidartinib, such as increase in plasma CSF-1 levels, and reduction in circulating nc-monocytes. These observations suggest clinically meaningful inhibition of CSF-1R as previously described *(26)*. These changes were however not as drastic as those observed with other CSF-1R inhibitors *(23)*. Despite this, we were unable to demonstrate reproducible decrease in intra-tumoral CD163^+^ TAMs in paired biopsies, although the actual number of analyzable pairs was low. Interestingly, one patient with PDAC who achieved 26% of tumor reduction had significant immune cell infiltration at baseline and we observed a reduction in TAMs and PMNs infiltration in the on-treatment sample. The initial tumor reduction was, however, not confirmed on subsequent scans which showed PD at week 16.

The reasons for the lack of clinical activity of pexidartinib combined with durvalumab remain unclear, but this observation is overall in line with recent reports of other studies of CSF-1R inhibitors combined with PD-(L)1 blockers *(37, 41, 42)*. Nevertheless, the activity observed in our study seems lower than that observed in the studies by Razak *et al. (37)* who observed PRs in 2/41 patients with MSS CRC (versus 0/18 in our study). In addition, we saw tumor shrinkage in patients with MSI-CRC with low exposure to pexidartinib (one patient was in the 400 mg/d cohort and one patient discontinued pexidartinib due AEs after less than 2 weeks). Our translational work points to alteration of the DC compartment by pexidartinib as a potential culprit. Indeed, we reported a sizeable decrease in absolute numbers of circulating DC subsets at C1D15 that resulted in decreased levels of IL-29/IFN-λ and to a lesser extent of IFN-α2a, respectively produced by cDC1 *(28)* and pDC *(27)*, in supernatants of short-term PBMC culture with TLR3/7/8-L. The activation of antitumor CD8^+^ T cells is a key step orchestrated by DC that depends on IFNs. pDC are specialized in IFN-I production in response to viral nucleic acids recognized by TLR 9 and 7 *(43)*. We and others have shown that loss of IFN-I production by dysfunctional tumor-associated pDC *(44–46)* is associated to regulatory T cells expansion *(47)*, worsening patient outcome in breast *(48)* and ovarian *(49)* cancers. In line with this observation, endogenous IFN-I was shown to play a central role in cancer immunosurveillance *via* the induction of specific CD8^+^ T cell responses *in vivo (33, 50)*. Antigen cross-presentation by XCR1^+^ cDC1 *(34, 51)* is also relevant for the induction and maintenance of antitumor CD8^+^ T cell responses. Hence, mouse XCR1^+^ cDC1 are necessary for spontaneous tumor rejection *(33, 34)* and efficiency of ICI *(35)*. Furthermore, XCR1^+^ cDC1 are unique in their capacity to produce large amounts of IFN-III (IFN-λ) in response to TLR3 ligands *(52)*. We previously identified IFN-λ-producing cDC1 in tumors and showed that high expression of XCR1^+^ cDC1 gene signature or *IFNL1* or its receptor is associated with improved prognosis in breast cancer *(28)*. Furthermore, the presence of IFN-λ in tumor supernatants correlated with high levels of CXCR3 ligands *(28)* that are essential for effector T cells recruitment *(53)*. All these observations converge toward a model where DC subsets and IFN-I/III act in synergy to promote an efficient immunity against cancer and led us to hypothesize that depletion of DC by pexidartinib may thus account for the lack of clinical activity of pexidartinib combined with anti-PD-L1.

CSF-1 is crucial in controlling macrophage differentiation, maintenance and proliferation *(12)*. In contrast, CSF-1 has only minor role in DC differentiation and homeostasis. Indeed, mice carrying *Csf1r*-null alleles show normal DC differentiation and numbers in peripheral lymphoid organs but have an impaired generation of non-lymphoid tissue DCs in the epidermis *(54)* and the lamina propria *(55)*, and of inflammatory DCs from monocytes *(56)*. The recent identification of the common DC precursor (CDP) that gives rise exclusively to pDC and cDC, but not monocytes *(57, 58)* established a DC-dedicated lineage, independent of that of macrophages and independent of CSF-1, in both rodents and humans. As we observed a global DC alteration in the blood of patients treated with pexidartinib, it is therefore unlikely that CSF-1 blockade is involved in this observation.

Pexidartinib was also shown to target the other FLT3 and SCF (c-KIT) RTK *(29)*, which are both known to play a major role in DC differentiation and survival (for review *(30)*). We observed a FLT3-L, but not SCF, increase in the plasma of patients treated with pexidartinib, suggesting a relevant impact of pexidartinib on FLT3 signaling (despite the differences in IC50 for CSF-1R and FLT3) that could explain the alterations of DC subset numbers observed in patients. In addition, we demonstrated that whereas pexidartinib did not affect GM-CSF-dependent BM-DC differentiation, it strongly inhibited the differentiation of all DC subsets in the presence of Flt3-L *in vitro*. pDCs and cDCs both develop from hematopoietic precursor cells under the influence of GM-CSF (for cDCs) and FLT3-L (for cDCs and pDCs). FLT3-L efficiently promotes the expansion of DCs and their precursors *in vivo* in human and mice *(59, 60)* and the differentiation of all DC subsets *in vitro (61)*. Consistently, lack of FLT3-L or its receptor FLT3 results in markedly reduced DC numbers *in vivo (59)*. Pexidartinib preferentially alters the early steps of DC differentiation from progenitors, but maintained activity at higher concentration on the survival of cDC. The impact on survival was not observed for human blood DC subsets suggesting their independence from FLT3-L when fully differentiated.

In this context, the use of mAbs to block CSF-1/CSF-1R would be preferable as it will not block other RTK, in combination with ICI. However, studies investigating CSF-1R-targeting mAbs (which do not inhibit FLT3 signaling) in combination with PD-1/PD-L1 blockers have shown only modest activity *(37, 41)*. Alternatively, depleting the nc-monocytes may not be sufficient to significantly impact TAM frequency and M2 polarization *in vivo*. In the study by Gomez-Roca *et al*., extensive biomarker work showed consistent depletion of CSF-1R/CD163^+^ TAMs, without significant modulation of T cell infiltration *(41)*. Some preclinical studies have shown that macrophage depletion could be rescued by myeloid-derived suppressor cells to maintain tumor-promoting immune suppression *(62)*.

Our finding that FLT3 inhibition with pexidartinib alters DC numbers may thus explain the failure of some TKI/ICI combinations in the clinic. This is a key finding, as many class-III TKI which are being explored as combination partner for PD-(L)1 blockers inhibit FLT3 in addition to their main clinical targets, and this is in particular the case for first generation VEGFR TKI such as sorafenib or sunitinib. In conclusion, our study on pexidartinib, provides new, clinically relevant information that may impact the interpretation of multiple on-going clinical trials combining TKI with ICI. The lack of activity of the TKI on FLT3 (and potentially other relevant RTK) should be ascertained before exploring combination with ICI in patients.

## MATERIALS AND METHODS

### Study design and patients’ selection

*Study design*. MEDIPLEX (NCT02777710) was a single arm, open label phase I study, designed to investigate the tolerability and preliminary efficacy of pexidartinib given orally twice daily (BID), combined with durvalumab at a fixed dose of 1500 mg, given intravenously (IV) every 4 weeks. The study comprised a dose escalation part and two expansion cohorts. In the dose escalation part, the dose of pexidartinib was escalated using a Likelihood Continual Reassessment Method (CRML) *(63)* with a 28-day window to evaluate the incidence of DLT (definitions provided in supplemental material), with the aim of defining a RP2D for pexidartinib combined with durvalumab. Once the RP2D was defined, 14 patients with PDAC and 14 patients with CRC were enrolled and treated at the RP2D in the extension part. The primary endpoint of the expansion part was the objective response rate according investigators after 16 weeks of treatment, and evaluated according to RECIST v1.1. For both parts, secondary endpoints included duration of response, progression free survival, overall survival and safety.

*Patients’ selection*. Adult patients, Eastern Cooperative Oncology Group performance status (ECOG PS) 0 or 1, with advanced/metastatic CRC and PDAC pretreated with at least one line of therapy and chemotherapy respectively for advanced disease were eligible. Adequate end-organ function, a RMH score of 0 or 1 *(64)* and a measurable lesion according to Response Evaluation Criteria in Solid Tumors version 1.1 (RECIST V1.1) were required. Additional inclusion criteria for patients enrolled in the expansion cohorts included: demonstration of at least one lesion amenable to repeat percutaneous sampling, and normal AST, ALT and bilirubin (reflecting the FDA requirements regarding pexidartinib after safety review of the ENLIVEN study *(39)*). All patients provided written informed consent. The study was approved by a central ethics committee and conducted in accordance with international standards of good clinical practice and all local laws and regulations.

### Study treatment

Pexidartinib was provided as 200 mg strength capsules and the initial dose of 400 mg/d (split into 200 mg in the morning and 200 mg in the evening) was selected based on pharmacodynamics data from prior human studies suggesting a PD effect at this dose. Pexidartinib was given orally and continuously and durvalumab was given IV on Day 1 of each 28-day cycle. Provisional dose levels were 400 (DL1), 600 (DL2), 800 (DL3), and 1000 (DL4) mg/d, as well as a -1 dose level at 400 mg/d 5 days on, 2 days off (intermittent schedule). Treatments were continued until disease progression, unacceptable toxicity, or patient withdrawal of consent.

### Tumor assessments and safety evaluations

Tumor measurements were assessed by imaging at baseline, every 8 weeks for the first 16 weeks, then every 12 weeks. Images were evaluated locally with use of the RECIST V1.1. Safety assessment included monitoring and recording of all AEs, regular safety lab tests and physical exams with monitoring of vital signs. AEs and laboratory abnormalities were graded according to the National Cancer Institute Common Terminology Criteria for Adverse Events (NCI-CTCAE, version 4.03).

### Pharmacokinetics

For all patients, plasma samples were collected at C1D1 and C1D15 at pre-dose and 1, 2, 4, and 6 hours post-Pexidartinib dosing (morning dose), then at pre-dose of C2D1 and C3D1. The AUC_0-6_ (Area under the concentration-time curve from time zero to 6 hours using linear-log trapezoidal rule) was calculated based on the plasma concentrations of PLX3397 according to the non-compartmental method *(65)*. PK calculations were performed using Phoenix WinNonlin 8.1 (Certara USA, Inc., Princeton, NJ).

### Cytokines quantification in plasma

The following cytokines were quantified in plasma of patients from either the dose escalation or the expansion using MSD technology according to the U-plex protocol from Mesoscale Discovery (MSD) and separated in 6 kits: CSF-1, IFN-α, IL-10, TNF-α, IFN-γ, IL-12p70, IL-29/IFNλ1, IL-1β, IL-6, IL-4, IL-13, IL-17A, IL-2, GM-CSF, SCF, and Flt3-L. Plasma were diluted at ½ before quantification.

### Phenotype and function of circulating immune cells

In expansion cohorts, blood samples were drawn at baseline (C1D1), during treatment (pre-dose of C1D15, C2D1, C3D1) and PBMC were generated on Ficoll gradient and frozen until their use. Frozen PBMC were then thawed, incubated for 2h at 37°C in complete Glutamax RPMI-1640 (Gibco) supplemented with 8% SAB (EFS), antibiotics (100 U/mL Penicillin, 100 μg/mL Streptomycin, Gibco) before experiments.

Characterization of innate and adaptive immune cell subsets was performed using multiparametric FC using specific surface marker combinations (see table S3). Cells were incubated with a viability marker FVS440UV (BD Biosciences) for 15 min at room temperature protected from light. Cells were washed and then stained with surface antibodies in the presence of BV stain buffer plus at 4°C for 30 min. After washes, cells were fixed in formaldehyde 4%. All FC acquisitions were done on a Fortessa-X20 Cell Analyzer (BD Biosciences), and data were processed in FlowJo 10.4 (Tree Star).

For functional analyses, 3×10^5^ PBMC were subjected to a short term (5 h) reactivation step in complete RPMI medium containing human sAB with TLR-L (R848 (TLR7/8) 5 μg/mL (InVivogen) or pI:C (TLR3), 30μg/mL (InVivogen)) or phorbol myristate acetate (PMA) (50ng/mL) /ionomycin (1μg/mL) (Sigma-Aldrich). Culture supernatants were collected and stored frozen until cytokine quantification by multiplex ECLIA (MSD) to analyze the functionality of cDC1 (IL-29 in response to pI:C), pDC (IFN-α in response to R848), monocytes (IL-1β, IL-6, TNFα in response to R848) and T cells (IL-2, IFN-γ, IL17-A, TNF-α, GM-CSF, IL-10 and IL-13 in response to PMA/ionomycin).

### Characterization of the tumor immune cell infiltrate by multiplex immunofluorescence

In expansion cohorts, patients consented to serial tumor biopsies (at baseline and at day 1 of cycle 2). 4 μm sections from formaldehyde-fixed and paraffin-embedded (FFPE) tumor biopsies were deparaffinized and a heat-induced antigen retrieval was performed in CC1 buffer for 36 min at 95°C. 3-4 colors multi-IF staining was performed with 4 panels of primary mAbs (Cytokeratin (CK)-MPO, CD3-CD20, CD68-CD163 and CD4-CD8-Ki67) and species-specific fluorochrome-coupled secondary mAbs (see table S4) on the DISCOVERY XT® autostainer (Ventana). The pool of primary mAbs appropriately diluted was incubated for 2 hours at 37°C then revealed by incubation for 1 hour at 37°C with the mix of fluorescently-labeled secondary mAbs. Sections were counterstained with Hoechst and mounted with a coverslip. Whole slides were imaged at a x20 magnification with the P150 Pannoramic scan-II FL (3D-Histech) and digital images were visualized with the CaseViewer software (3D-Histech).

### *In vitro* differentiation of murine BM-DC

Tibias and femurs from C57Bl/6 mice were removed under sterile conditions. Both ends of the bone were cut and BM cells were flushed with 5mL of PBS and centrifuged at 1300rpm for 8 min at 4°C. Red blood cells were lysed in 1mL of BD Pharmlyse for 1min at room temperature. Cells were centrifuged at 1300rpm for 8 min at 4°C, washed with 10mL of PBS, and passed on 30μm filter. BM cells were then resuspended in RPMI-1640 medium supplemented with 10% Fetal Calf Serum (Eurobio), β-mercaptoethanol (0.1%), Penicillin/streptomycin, L-Glutamine (200 μM), non-essential amino acids and Hepes (1mM) (all from Gibco) and plated into 24-well plates at a density of 2×10^6^ cell/mL with recombinant Flt3-L (rFlt3-L; 50ng/mL, Miltenyi Biotec), B16-Flt3-L culture supernatant (sFlt3-L, 5%); or GM-CSF (25ng/mL, Peprotech). Cells were cultured at 37°C, 5% CO2 with or without different doses of pexidartinib (0.01μM to 40μM). For rFlt3-L- and GM-CSF conditions, half of the culture medium was changed at day 3. At day 7, cells were collected and DC subsets were identified by FC using panel 3 (table S3).

### Analysis of human DC survival

Healthy donor (HD) blood samples were obtained from the *Etablissement Français du Sang* (EFS, Lyon) (convention #19-023). PBMCs were isolated by ficoll from fresh HD whole blood. They were cultured 18 hours a 37°C, 5% CO2 in complete RPMI supplemented with 10% FCS with or without graded doses of pexidartinib (1μM, 10μM and 40μM). Cells were then collected, centrifuged at 1400rpm for 4 min at 4°C and washed in PBS. Then they were resuspended in 100μL of staining buffer and incubated at 4°C for 30 min protected from light with surface antibodies according to panel 4 (table S3) in the presence of BV stain buffer plus and LD-Zombie-Near-IR as viability marker. Cells were washed again and fixed with formaldehyde 4%. Viable DC subsets were identified by FC as previously described *(27)*. All FC acquisitions were done on a Fortessa-X20 Cell Analyzer, and data were processed in FlowJo 10.8.

### Statistical analysis

#### Sample size

The dose escalation scheme was done using a CRML: successive cohorts of 3 patients treated with the combination therapy. The sample size for the expansion cohorts was calculated according to the first stage of Gehan design: 14 patients per cohort allowed for eliminating a treatment with response rate lower than 20%. Indeed, if no clinical responses are observed among 14 patients, the probability that the RR is greater than 20% will be less than 5%.

#### Analysis

CRM analyses were performed using R® v3.3.1 statistical software and clinical data analyses using SAS® v9.4 statistical software. Kaplan-Meier method was used to estimate OS and PFS. Biological data statistical calculations were performed in GraphPad Prism 9. A one-way ANOVA (Kruskal-Wallis test) followed by Dunn’s multiple comparisons to adjust for false discovery rate or a student’s *t*-test was used for human experiments. A one-way ANOVA followed by Dunnett’s multiple comparisons test or a 2-way ANOVA followed by Bonferroni’s multiple comparisons test was used for mouse *in vitro* analysis. Differences were considered significant at P < 0.05.

## Data Availability

All data produced in the present study are available upon reasonable request to the authors

## Acknowledgments

The MEDIPLEX trial was sponsored by Centre Léon Bérard. We thank the patients and their families for participating in this study. We thank the investigators, nurses, radiology and pharmacy teams and other study staff for their contributions to this trial. We also wish to thank the CRB Centre Léon Bérard, Lyon France [BB-0033-00050] for logistic and preparation of biological samples, A Vermorel and L Odeyer for selection of biopsy tumor samples and technical multi-immunofluorescence. Mouse experiments were performed at the P-PAC animal facility of the CRCL.

## Author contribution

P.A.C., G.G., A-S.B., S.C., L.M., and D.P. conceived and designed the clinical study. S.C., L.M., and D.P. developed the methodology. P.A.C., C.G.R., L.E., C.T., I.K., and J-P.D. managed patients’ cohorts. C.C., N.B-V., C.M-C., B.D., A.V., C.R., and I.T. conceived and designed the ancillary study. P.A.C., C.G.R., A.V., L.E., C.T., I.K., J-P.D., C.R., J.B., A.N’K., and I.T. acquired the data. P.A.C., S.C., L.M., N.B-V., B.D., C.M-C., C.R., A.N’K., A.V., and J.B. analyzed and interpreted the data. C.M-C., P.A.C., N.B-V., C.C., B.D., A.V., C.R., and A-S.B. wrote and edited the paper. P.A.C., G.G., C.M-C., C.C., and N.B-V. supervised the study.

## Data and materials availability

All data associated with this study are present in the paper or the Supplementary Materials.

## REFERENCES

1. L. Rahib, B. D. Smith, R. Aizenberg, A. B. Rosenzweig, J. M. Fleshman, L. M. Matrisian, Projecting cancer incidence and deaths to 2030: the unexpected burden of thyroid, liver, and pancreas cancers in the United States. Cancer Res 74, 2913–2921 (2014).

2. E. Dekker, P. J. Tanis, J. L. A. Vleugels, P. M. Kasi, M. B. Wallace, Colorectal cancer. Lancet 394, 1467–1480 (2019).

3. J. D. Mizrahi, R. Surana, J. W. Valle, R. T. Shroff, Pancreatic cancer. Lancet 395, 2008–2020 (2020).

4. J. R. Brahmer, S. S. Tykodi, L. Q. M. Chow, W.-J. Hwu, S. L. Topalian, P. Hwu, C. G. Drake, L. H. Camacho, J. Kauh, K. Odunsi, H. C. Pitot, O. Hamid, S. Bhatia, R. Martins, K. Eaton, S. Chen, T. M. Salay, S. Alaparthy, J. F. Grosso, A. J. Korman, S. M. Parker, S. Agrawal, S. M. Goldberg, D. M. Pardoll, A. Gupta, J. M. Wigginton, Safety and activity of anti-PD-L1 antibody in patients with advanced cancer. N Engl J Med 366, 2455–2465 (2012).

5. D. T. Le, J. N. Durham, K. N. Smith, H. Wang, B. R. Bartlett, L. K. Aulakh, S. Lu, H. Kemberling, C. Wilt, B. S. Luber, F. Wong, N. S. Azad, A. A. Rucki, D. Laheru, R. Donehower, A. Zaheer, G. A. Fisher, T. S. Crocenzi, J. J. Lee, T. F. Greten, A. G. Duffy, K. K. Ciombor, A. D. Eyring, B. H. Lam, A. Joe, S. P. Kang, M. Holdhoff, L. Danilova, L. Cope, C. Meyer, S. Zhou, R. M. Goldberg, D. K. Armstrong, K. M. Bever, A. N. Fader, J. Taube, F. Housseau, D. Spetzler, N. Xiao, D. M. Pardoll, N. Papadopoulos, K. W. Kinzler, J. R. Eshleman, B. Vogelstein, R. A. Anders, L. A. Diaz, Mismatch repair deficiency predicts response of solid tumors to PD-1 blockade. Science 357, 409–413 (2017).

6. E. M. O’Reilly, D.-Y. Oh, N. Dhani, D. J. Renouf, M. A. Lee, W. Sun, G. Fisher, A. Hezel, S.-C. Chang, G. Vlahovic, O. Takahashi, Y. Yang, D. Fitts, P. A. Philip, Durvalumab With or Without Tremelimumab for Patients With Metastatic Pancreatic Ductal Adenocarcinoma: A Phase 2 Randomized Clinical Trial. JAMA Oncol 5, 1431–1438 (2019).

7. R. A. Droeser, C. Hirt, C. T. Viehl, D. M. Frey, C. Nebiker, X. Huber, I. Zlobec, S. Eppenberger-Castori, Tzankov, R. Rosso, M. Zuber, M. G. Muraro, F. Amicarella, E. Cremonesi, M. Heberer, G. Iezzi, A. Lugli, L. Terracciano, G. Sconocchia, D. Oertli, G. C. Spagnoli, L. Tornillo, Clinical impact of programmed cell death ligand 1 expression in colorectal cancer. Eur J Cancer 49, 2233–2242 (2013).

8. L. Danilova, W. J. Ho, Q. Zhu, T. Vithayathil, A. De Jesus-Acosta, N. S. Azad, D. A. Laheru, E. J. Fertig, R. Anders, E. M. Jaffee, M. Yarchoan, Programmed Cell Death Ligand-1 (PD-L1) and CD8 Expression Profiling Identify an Immunologic Subtype of Pancreatic Ductal Adenocarcinomas with Favorable Survival. Cancer Immunol Res 7, 886–895 (2019).

9. K. Ganesh, Z. K. Stadler, A. Cercek, R. B. Mendelsohn, J. Shia, N. H. Segal, L. A. Diaz, Immunotherapy in colorectal cancer: rationale, challenges and potential. Nat Rev Gastroenterol Hepatol 16, 361–375 (2019).

10. W. J. Ho, E. M. Jaffee, L. Zheng, The tumour microenvironment in pancreatic cancer - clinical challenges and opportunities. Nat Rev Clin Oncol 17, 527–540 (2020).

11. R. N. Ramos, C. Rodriguez, M. Hubert, M. Ardin, I. Treilleux, C. H. Ries, E. Lavergne, S. Chabaud, A. Colombe, O. Trédan, H. G. Guedes, F. Laginha, W. Richer, E. Piaggio, J. A. M. Barbuto, C. Caux, C. Ménétrier-Caux, N. Bendriss-Vermare, CD163+ tumor-associated macrophage accumulation in breast cancer patients reflects both local differentiation signals and systemic skewing of monocytes. Clin Transl Immunology 9, e1108 (2020).

12. I. Ushach, A. Zlotnik, Biological role of granulocyte macrophage colony-stimulating factor (GM-CSF) and macrophage colony-stimulating factor (M-CSF) on cells of the myeloid lineage. J Leukoc Biol 100, 481–489 (2016).

13. A. Freuchet, A. Salama, S. Remy, C. Guillonneau, I. Anegon, IL-34 and CSF-1, deciphering similarities and differences at steady state and in diseases. J Leukoc Biol 110, 771–796 (2021).

14. D. Laoui, E. Van Overmeire, P. De Baetselier, J. A. Van Ginderachter, G. Raes, Functional Relationship between Tumor-Associated Macrophages and Macrophage Colony-Stimulating Factor as Contributors to Cancer Progression. Front Immunol 5, 489 (2014).

15. E. Van Overmeire, B. Stijlemans, F. Heymann, J. Keirsse, Y. Morias, Y. Elkrim, L. Brys, C. Abels, Q. Lahmar, C. Ergen, L. Vereecke, F. Tacke, P. De Baetselier, J. A. Van Ginderachter, D. Laoui, M-CSF and GM-CSF Receptor Signaling Differentially Regulate Monocyte Maturation and Macrophage Polarization in the Tumor Microenvironment. Cancer Res 76, 35–42 (2016).

16. Y. Zhu, B. L. Knolhoff, M. A. Meyer, T. M. Nywening, B. L. West, J. Luo, A. Wang-Gillam, S. P. Goedegebuure, D. C. Linehan, D. G. DeNardo, CSF1/CSF1R blockade reprograms tumor-infiltrating macrophages and improves response to T-cell checkpoint immunotherapy in pancreatic cancer models. Cancer Res 74, 5057–5069 (2014).

17. E. Peranzoni, J. Lemoine, L. Vimeux, V. Feuillet, S. Barrin, C. Kantari-Mimoun, N. Bercovici, M. Guérin, J. Biton, H. Ouakrim, F. Régnier, A. Lupo, M. Alifano, D. Damotte, E. Donnadieu, Macrophages impede CD8 T cells from reaching tumor cells and limit the efficacy of anti-PD-1 treatment. Proc Natl Acad Sci U S A 115, E4041–E4050 (2018).

18. M. A. Cannarile, M. Weisser, W. Jacob, A.-M. Jegg, C. H. Ries, D. Rüttinger, Colony-stimulating factor 1 receptor (CSF1R) inhibitors in cancer therapy. J Immunother Cancer 5, 53 (2017).

19. C. H. Ries, S. Hoves, M. A. Cannarile, D. Rüttinger, CSF-1/CSF-1R targeting agents in clinical development for cancer therapy. Curr Opin Pharmacol 23, 45–51 (2015).

20. W. D. Tap, Z. A. Wainberg, S. P. Anthony, P. N. Ibrahim, C. Zhang, J. H. Healey, B. Chmielowski, A. P. Staddon, A. L. Cohn, G. I. Shapiro, V. L. Keedy, A. S. Singh, I. Puzanov, E. L. Kwak, A. J. Wagner, D. D. Von Hoff, G. J. Weiss, R. K. Ramanathan, J. Zhang, G. Habets, Y. Zhang, E. A. Burton, G. Visor, L. Sanftner, P. Severson, H. Nguyen, M. J. Kim, A. Marimuthu, G. Tsang, R. Shellooe, C. Gee, B. L. West, P. Hirth, K. Nolop, M. van de Rijn, H. H. Hsu, C. Peterfy, P. S. Lin, S. Tong-Starksen, G. Bollag, Structure-Guided Blockade of CSF1R Kinase in Tenosynovial Giant-Cell Tumor. N Engl J Med 373, 428–437 (2015).

21. J.-H. Lee, T. W.-W. Chen, C.-H. Hsu, Y.-H. Yen, J. C.-H. Yang, A.-L. Cheng, S.-I. Sasaki, L. L. Chiu, M. Sugihara, T. Ishizuka, T. Oguma, N. Tajima, C.-C. Lin, A phase I study of pexidartinib, a colony-stimulating factor 1 receptor inhibitor, in Asian patients with advanced solid tumors. Invest New Drugs 38, 99–110 (2020).

22. N. Butowski, H. Colman, J. F. De Groot, A. M. Omuro, L. Nayak, P. Y. Wen, T. F. Cloughesy, A. Marimuthu, S. Haidar, A. Perry, J. Huse, J. Phillips, B. L. West, K. B. Nolop, H. H. Hsu, K. L. Ligon, A. M. Molinaro, M. Prados, Orally administered colony stimulating factor 1 receptor inhibitor PLX3397 in recurrent glioblastoma: an Ivy Foundation Early Phase Clinical Trials Consortium phase II study. Neuro Oncol 18, 557–564 (2016).

23. P. A. Cassier, A. Italiano, C. A. Gomez-Roca, C. Le Tourneau, M. Toulmonde, M. A. Cannarile, C. Ries, Brillouet, C. Müller, A.-M. Jegg, A.-M. Bröske, M. Dembowski, K. Bray-French, C. Freilinger, G. Meneses-Lorente, M. Baehner, R. Harding, J. Ratnayake, K. Abiraj, N. Gass, K. Noh, R. D. Christen, L. Ukarma, E. Bompas, J.-P. Delord, J.-Y. Blay, D. Rüttinger, CSF1R inhibition with emactuzumab in locally advanced diffuse-type tenosynovial giant cell tumours of the soft tissue: a dose-escalation and dose-expansion phase 1 study. Lancet Oncol 16, 949–956 (2015).

24. C. C. Smith, C. Zhang, K. C. Lin, E. A. Lasater, Y. Zhang, E. Massi, L. E. Damon, M. Pendleton, A. Bashir, R. Sebra, A. Perl, A. Kasarskis, R. Shellooe, G. Tsang, H. Carias, B. Powell, E. A. Burton, B. Matusow, J. Zhang, W. Spevak, P. N. Ibrahim, M. H. Le, H. H. Hsu, G. Habets, B. L. West, G. Bollag, N. P. Shah, Characterizing and Overriding the Structural Mechanism of the Quizartinib-Resistant FLT3 “Gatekeeper” F691L Mutation with PLX3397. Cancer Discov 5, 668–679 (2015).

25. C. H. Ries, M. A. Cannarile, S. Hoves, J. Benz, K. Wartha, V. Runza, F. Rey-Giraud, L. P. Pradel, F. Feuerhake, I. Klaman, T. Jones, U. Jucknischke, S. Scheiblich, K. Kaluza, I. H. Gorr, A. Walz, K. Abiraj, P. A. Cassier, A. Sica, C. Gomez-Roca, K. E. de Visser, A. Italiano, C. Le Tourneau, J.-P. Delord, H. Levitsky, J.-Y. Blay, D. Rüttinger, Targeting tumor-associated macrophages with anti-CSF-1R antibody reveals a strategy for cancer therapy. Cancer Cell 25, 846–859 (2014).

26. R. Wesolowski, N. Sharma, L. Reebel, M. B. Rodal, A. Peck, B. L. West, A. Marimuthu, P. Severson, D. A. Karlin, A. Dowlati, M. H. Le, L. M. Coussens, H. S. Rugo, Phase Ib study of the combination of pexidartinib (PLX3397), a CSF-1R inhibitor, and paclitaxel in patients with advanced solid tumors. Ther Adv Med Oncol 11, 1758835919854238 (2019).

27. E. Verronèse, A. Delgado, J. Valladeau-Guilemond, G. Garin, S. Guillemaut, O. Tredan, I. Ray-Coquard, T. Bachelot, A. N’Kodia, C. Bardin-Dit-Courageot, C. Rigal, D. Pérol, C. Caux, C. Ménétrier-Caux, Immune cell dysfunctions in breast cancer patients detected through whole blood multi-parametric flow cytometry assay. Oncoimmunology 5, e1100791 (2016).

28. M. Hubert, E. Gobbini, C. Couillault, T.-P. V. Manh, A.-C. Doffin, J. Berthet, C. Rodriguez, V. Ollion, J. Kielbassa, C. Sajous, I. Treilleux, O. Tredan, B. Dubois, M. Dalod, N. Bendriss-Vermare, C. Caux, J. Valladeau-Guilemond, IFN-III is selectively produced by cDC1 and predicts good clinical outcome in breast cancer. Sci Immunol 5, eaav3942 (2020).

29. Y. N. Lamb, Pexidartinib: First Approval. Drugs 79, 1805–1812 (2019).

30. F. J. Cueto, D. Sancho, The Flt3L/Flt3 Axis in Dendritic Cell Biology and Cancer Immunotherapy. Cancers (Basel) 13, 1525 (2021).

31. K. Brasel, T. De Smedt, J. L. Smith, C. R. Maliszewski, Generation of murine dendritic cells from flt3-ligand-supplemented bone marrow cultures. Blood 96, 3029–3039 (2000).

32. K. Inaba, M. Inaba, N. Romani, H. Aya, M. Deguchi, S. Ikehara, S. Muramatsu, R. M. Steinman, Generation of large numbers of dendritic cells from mouse bone marrow cultures supplemented with granulocyte/macrophage colony-stimulating factor. J Exp Med 176, 1693–1702 (1992).

33. R. Mattiuz, C. Brousse, M. Ambrosini, J.-C. Cancel, G. Bessou, J. Mussard, A. Sanlaville, C. Caux, N. Bendriss-Vermare, J. Valladeau-Guilemond, M. Dalod, K. Crozat, Type 1 conventional dendritic cells and interferons are required for spontaneous CD4+ and CD8+ T-cell protective responses to breast cancer. Clin Transl Immunology 10, e1305 (2021).

34. K. Hildner, B. T. Edelson, W. E. Purtha, M. Diamond, H. Matsushita, M. Kohyama, B. Calderon, B. U. Schraml, E. R. Unanue, M. S. Diamond, R. D. Schreiber, T. L. Murphy, K. M. Murphy, Batf3 deficiency reveals a critical role for CD8alpha+ dendritic cells in cytotoxic T cell immunity. Science 322, 1097–1100 (2008).

35. H. Salmon, J. Idoyaga, A. Rahman, M. Leboeuf, R. Remark, S. Jordan, M. Casanova-Acebes, M. Khudoynazarova, J. Agudo, N. Tung, S. Chakarov, C. Rivera, B. Hogstad, M. Bosenberg, D. Hashimoto, S. Gnjatic, N. Bhardwaj, A. K. Palucka, B. D. Brown, J. Brody, F. Ginhoux, M. Merad, Expansion and Activation of CD103(+) Dendritic Cell Progenitors at the Tumor Site Enhances Tumor Responses to Therapeutic PD-L1 and BRAF Inhibition. Immunity 44, 924–938 (2016).

36. Z. A. Radi, P. H. Koza-Taylor, R. R. Bell, L. A. Obert, H. A. Runnels, J. S. Beebe, M. P. Lawton, S. Sadis, Increased serum enzyme levels associated with kupffer cell reduction with no signs of hepatic or skeletal muscle injury. Am J Pathol 179, 240–247 (2011).

37. A. R. Razak, J. M. Cleary, V. Moreno, M. Boyer, E. Calvo Aller, W. Edenfield, J. Tie, R. D. Harvey, A. Rutten, M. A. Shah, A. J. Olszanski, D. Jäger, N. Lakhani, D. P. Ryan, E. Rasmussen, G. Juan, H. Wong, N. Soman, M.-A. D. Smit, D. Nagorsen, K. P. Papadopoulos, Safety and efficacy of AMG 820, an anti-colony-stimulating factor 1 receptor antibody, in combination with pembrolizumab in adults with advanced solid tumors. J Immunother Cancer 8, e001006 (2020).

38. S. Bissinger, C. Hage, V. Wagner, I.-P. Maser, V. Brand, M. Schmittnaegel, A.-M. Jegg, M. Cannarile, C. Watson, I. Klaman, N. Rieder, A. González Loyola, T. V. Petrova, P. A. Cassier, C. Gomez-Roca, V. Sibaud, M. De Palma, S. Hoves, C. H. Ries, Macrophage depletion induces edema through release of matrix-degrading proteases and proteoglycan deposition. Sci Transl Med 13, eabd4550 (2021).

39. W. D. Tap, H. Gelderblom, E. Palmerini, J. Desai, S. Bauer, J.-Y. Blay, T. Alcindor, K. Ganjoo, J. Martín-Broto, C. W. Ryan, D. M. Thomas, C. Peterfy, J. H. Healey, M. van de Sande, H. L. Gelhorn, D. E. Shuster, Q. Wang, A. Yver, H. H. Hsu, P. S. Lin, S. Tong-Starksen, S. Stacchiotti, A. J. Wagner, ENLIVEN investigators, Pexidartinib versus placebo for advanced tenosynovial giant cell tumour (ENLIVEN): a randomised phase 3 trial. Lancet 394, 478–487 (2019).

40. S. J. Antonia, A. Villegas, D. Daniel, D. Vicente, S. Murakami, R. Hui, T. Yokoi, A. Chiappori, K. H. Lee, M. de Wit, B. C. Cho, M. Bourhaba, X. Quantin, T. Tokito, T. Mekhail, D. Planchard, Y.-C. Kim, C. S. Karapetis, S. Hiret, G. Ostoros, K. Kubota, J. E. Gray, L. Paz-Ares, J. de Castro Carpeño, C. Wadsworth, G. Melillo, H. Jiang, Y. Huang, P. A. Dennis, M. Özgüroğlu, PACIFIC Investigators, Durvalumab after Chemoradiotherapy in Stage III Non-Small-Cell Lung Cancer. N Engl J Med 377, 1919–1929 (2017).

41. C. Gomez-Roca, P. A. Cassier, D. Zamarin, J.-P. Machiels, J. L. Perez-Gracia, F. S. Hodi, A. Taus, M. Martinez Garcia, V. Boni, J. Eder, N. Hafez, R. J. Sullivan, D. F. McDermott, S. Champiat, S. Aspeslagh, C. Terret, A.-M. Jegg, W. Jacob, M. A. Cannarile, C. H. Ries, K. Korski, F. Michielin, R. D. Christen, G. Babitzki, C. Watson, G. Meneses-Lorente, M. Weisser, D. Rüttinger, J.-P. Delord, A. Marabelle, Anti-CSF-1R emactuzumab in combination with anti-PD-L1 atezolizumab in advanced solid tumor patients naïve or experienced for immune checkpoint blockadeJournal for ImmunoTherapy of Cancer (in press).

42. M. Johnson, A. Z. Dudek, A. Sukari, J. Call, P. R. Kunk, K. Lewis, J. F. Gainor, J. Sarantopoulos, P. Lee, Golden, A. Harney, S. M. Rothenberg, Y. Zhang, J. W. Goldman, ARRY-382 in Combination with Pembrolizumab in Patients with Advanced Solid Tumors: Results from a Phase 1b/2 Study. Clin Cancer Res 28, 2517–2526 (2022).

43. M. Swiecki, M. Colonna, The multifaceted biology of plasmacytoid dendritic cells. Nat Rev Immunol 15, 471–485 (2015).

44. V. Sisirak, J. Faget, M. Gobert, N. Goutagny, N. Vey, I. Treilleux, S. Renaudineau, G. Poyet, S. I. Labidi-Galy, S. Goddard-Leon, I. Durand, I. Le Mercier, A. Bajard, T. Bachelot, A. Puisieux, I. Puisieux, J.-Y. Blay, C. Ménétrier-Caux, C. Caux, N. Bendriss-Vermare, Impaired IFN-α production by plasmacytoid dendritic cells favors regulatory T-cell expansion that may contribute to breast cancer progression. Cancer Res 72, 5188–5197 (2012).

45. S. I. Labidi-Galy, V. Sisirak, P. Meeus, M. Gobert, I. Treilleux, A. Bajard, J.-D. Combes, J. Faget, F. Mithieux, A. Cassignol, O. Tredan, I. Durand, C. Ménétrier-Caux, C. Caux, J.-Y. Blay, I. Ray-Coquard, N. Bendriss-Vermare, Quantitative and functional alterations of plasmacytoid dendritic cells contribute to immune tolerance in ovarian cancer. Cancer Res 71, 5423–5434 (2011).

46. E. Hartmann, B. Wollenberg, S. Rothenfusser, M. Wagner, D. Wellisch, B. Mack, T. Giese, O. Gires, S. Endres, G. Hartmann, Identification and functional analysis of tumor-infiltrating plasmacytoid dendritic cells in head and neck cancer. Cancer Res 63, 6478–6487 (2003).

47. J. Faget, N. Bendriss-Vermare, M. Gobert, I. Durand, D. Olive, C. Biota, T. Bachelot, I. Treilleux, S. Goddard-Leon, E. Lavergne, S. Chabaud, J. Y. Blay, C. Caux, C. Ménétrier-Caux, ICOS-ligand expression on plasmacytoid dendritic cells supports breast cancer progression by promoting the accumulation of immunosuppressive CD4+ T cells. Cancer Res 72, 6130–6141 (2012).

48. I. Treilleux, J.-Y. Blay, N. Bendriss-Vermare, I. Ray-Coquard, T. Bachelot, J.-P. Guastalla, A. Bremond, S. Goddard, J.-J. Pin, C. Barthelemy-Dubois, S. Lebecque, Dendritic cell infiltration and prognosis of early stage breast cancer. Clin Cancer Res 10, 7466–7474 (2004).

49. S. I. Labidi-Galy, I. Treilleux, S. Goddard-Leon, J.-D. Combes, J.-Y. Blay, I. Ray-Coquard, C. Caux, N. Bendriss-Vermare, Plasmacytoid dendritic cells infiltrating ovarian cancer are associated with poor prognosis. Oncoimmunology 1, 380–382 (2012).

50. M. S. Diamond, M. Kinder, H. Matsushita, M. Mashayekhi, G. P. Dunn, J. M. Archambault, H. Lee, C. D. Arthur, J. M. White, U. Kalinke, K. M. Murphy, R. D. Schreiber, Type I interferon is selectively required by dendritic cells for immune rejection of tumors. J Exp Med 208, 1989–2003 (2011).

51. S. L. Jongbloed, A. J. Kassianos, K. J. McDonald, G. J. Clark, X. Ju, C. E. Angel, C.-J. J. Chen, P. R. Dunbar, R. B. Wadley, V. Jeet, A. J. E. Vulink, D. N. J. Hart, K. J. Radford, Human CD141+ (BDCA-3)+ dendritic cells (DCs) represent a unique myeloid DC subset that cross-presents necrotic cell antigens. Journal of Experimental Medicine 207, 1247–1260 (2010).

52. H. Lauterbach, B. Bathke, S. Gilles, C. Traidl-Hoffmann, C. A. Luber, G. Fejer, M. A. Freudenberg, G. M. Davey, D. Vremec, A. Kallies, L. Wu, K. Shortman, P. Chaplin, M. Suter, M. O’Keeffe, H. Hochrein, Mouse CD8α+ DCs and human BDCA3+ DCs are major producers of IFN-λ in response to poly IC. Journal of Experimental Medicine 207, 2703–2717 (2010).

53. M. T. Chow, A. J. Ozga, R. L. Servis, D. T. Frederick, J. A. Lo, D. E. Fisher, G. J. Freeman, G. M. Boland, A. D. Luster, Intratumoral Activity of the CXCR3 Chemokine System Is Required for the Efficacy of Anti-PD-1 Therapy. Immunity 50, 1498–1512.e5 (2019).

54. F. Ginhoux, F. Tacke, V. Angeli, M. Bogunovic, M. Loubeau, X.-M. Dai, E. R. Stanley, G. J. Randolph, M. Merad, Langerhans cells arise from monocytes in vivo. Nat Immunol 7, 265–273 (2006).

55. M. Bogunovic, F. Ginhoux, J. Helft, L. Shang, D. Hashimoto, M. Greter, K. Liu, C. Jakubzick, M. A. Ingersoll, M. Leboeuf, E. R. Stanley, M. Nussenzweig, S. A. Lira, G. J. Randolph, M. Merad, Origin of the lamina propria dendritic cell network. Immunity 31, 513–525 (2009).

56. M. Greter, J. Helft, A. Chow, D. Hashimoto, A. Mortha, J. Agudo-Cantero, M. Bogunovic, E. L. Gautier, J. Miller, M. Leboeuf, G. Lu, C. Aloman, B. D. Brown, J. W. Pollard, H. Xiong, G. J. Randolph, J. E. Chipuk, P. S. Frenette, M. Merad, GM-CSF controls nonlymphoid tissue dendritic cell homeostasis but is dispensable for the differentiation of inflammatory dendritic cells. Immunity 36, 1031–1046 (2012).

57. J. Lee, G. Breton, T. Y. K. Oliveira, Y. J. Zhou, A. Aljoufi, S. Puhr, M. J. Cameron, R.-P. Sékaly, M. C. Nussenzweig, K. Liu, Restricted dendritic cell and monocyte progenitors in human cord blood and bone marrow. J Exp Med 212, 385–399 (2015).

58. K. Liu, G. D. Victora, T. A. Schwickert, P. Guermonprez, M. M. Meredith, K. Yao, F.-F. Chu, G. J. Randolph, A. Y. Rudensky, M. Nussenzweig, In vivo analysis of dendritic cell development and homeostasis. Science 324, 392–397 (2009).

59. H. J. McKenna, K. L. Stocking, R. E. Miller, K. Brasel, T. De Smedt, E. Maraskovsky, C. R. Maliszewski, D. H. Lynch, J. Smith, B. Pulendran, E. R. Roux, M. Teepe, S. D. Lyman, J. J. Peschon, Mice lacking flt3 ligand have deficient hematopoiesis affecting hematopoietic progenitor cells, dendritic cells, and natural killer cells. Blood 95, 3489–3497 (2000).

60. B. Pulendran, J. Banchereau, S. Burkeholder, E. Kraus, E. Guinet, C. Chalouni, D. Caron, C. Maliszewski, J. Davoust, J. Fay, K. Palucka, Flt3-ligand and granulocyte colony-stimulating factor mobilize distinct human dendritic cell subsets in vivo. J Immunol 165, 566–572 (2000).

61. S. H. Naik, A. I. Proietto, N. S. Wilson, A. Dakic, P. Schnorrer, M. Fuchsberger, M. H. Lahoud, M. O’Keeffe, Q. Shao, W. Chen, J. A. Villadangos, K. Shortman, L. Wu, Cutting edge: generation of splenic CD8+ and CD8-dendritic cell equivalents in Fms-like tyrosine kinase 3 ligand bone marrow cultures. J Immunol 174, 6592–6597 (2005).

62. T. M. Nywening, B. A. Belt, D. R. Cullinan, R. Z. Panni, B. J. Han, D. E. Sanford, R. C. Jacobs, J. Ye, A. A. Patel, W. E. Gillanders, R. C. Fields, D. G. DeNardo, W. G. Hawkins, P. Goedegebuure, D. C. Linehan, Targeting both tumour-associated CXCR2+ neutrophils and CCR2+ macrophages disrupts myeloid recruitment and improves chemotherapeutic responses in pancreatic ductal adenocarcinoma. Gut 67, 1112–1123 (2018).

63. S. M. Lee, null Ying Kuen Cheung, Model calibration in the continual reassessment method. Clin Trials 6, 227–238 (2009).

64. H.-T. Arkenau, J. Barriuso, D. Olmos, J. E. Ang, J. de Bono, I. Judson, S. Kaye, Prospective validation of a prognostic score to improve patient selection for oncology phase I trials. J Clin Oncol 27, 2692–2696 (2009).

65. M. Gibaldi, D. Perrier, Eds., Pharmacokinetics (CRC Press, ed. 0, 1982; https://www.taylorfrancis.com/books/9781420089141).

