## supplemental material for "The CSF-1R inhibitor Pexidartinib impacts dendritic cell differentiation through inhibition of FLT3 signaling and may antagonize the effect of durvalumab in patients with advanced cancer – results from a phase 1 study"

### SUPPLEMENTARY MATERIALS

**DLT definition:** DLTs were defined as: any Grade ≥4 immune related adverse event (irAE), Any Grade ≥3 irAE, that does not downgrade to Grade 2 within 3 days after onset of the event despite optimal medical management including systemic corticosteroids or does not downgrade to ≤ Grade 1 or baseline within 14 days; Grade 2 pneumonitis that does not resolve to ≤ Grade 1 within 3 days of the initiation of maximal supportive care; Grade ≥ 4 neutropenia (ANC < 500/µL) lasting ≥ 7 days, Grade ≥ 3 febrile neutropenia, Grade ≥ 4 thrombocytopenia associated with clinically significant bleeding and lasting > 72 hours; liver transaminase > 10 x ULN, AST/ALT > 3x ULN with concurrent bilirubin >2x ULN; any other study drug related toxicity considered significant enough to be qualified as DLT in the opinion of the investigators after discussion with the sponsor; Study-drug related toxicity leading to administration of less than 80% of the planned dose of pexidartinib during the first cycle of therapy); Any other Grade ≥3 major organ adverse events with exception.

**DLT exceptions:** Grade 3 fatigue lasting ≤ 7 days; Grade 3 endocrine disorder (thyroid, pituitary, and/or adrenal insufficiency) that is managed with or without systemic corticosteroid therapy and/or hormone replacement therapy and the patient is asymptomatic ; Grade 3 inflammatory reaction attributed to a local antitumor response (eg, inflammatory reaction at sites of metastatic disease, lymph nodes, etc); Grade 3 infusion-related reaction (first occurrence and in the absence of steroid prophylaxis) that resolves within 6 hours with appropriate clinical management; Grade 3 neutropenia that is not associated with fever or systemic infection that improves by at least 1 grade within 7 days. Grade 3 or Grade 4 febrile neutropenia will be a DLT regardless of duration or reversibility; Grade 3 or 4 lymphopenia; Grade 3 thrombocytopenia that is not associated with clinically significant bleeding that requires medical intervention, and improves by at least 1 grade within 3 days; Isolated Grade 3 electrolyte abnormalities that are not associated with clinical signs or symptoms and are reversed with appropriate maximal medical intervention within 3 days.

### SUPPLEMENTARY FIGURE LEGENDS AND TABLES

**Supplementary Fig. 1: Progression-Free Survival (PFS) and Overall Survival (OS).** Kaplan–Meier curves by RECIST version 1.1 for **(A)** PFS and **(B)** OS in 28 evaluable patients (expansion part) with advanced/metastatic CRC and PDAC. Median PFS and OS are indicated in months. Number of patients at risk at indicated time points are shown below the x-axis.

**Supplementary Fig. 2: Persistency of high CSF-1 levels only in patients who continued to receive pexidartinib.** CSF-1 levels were quantified by multiplex ECLIA in plasma of patients who have continuously taken pexidartinib versus those who have stopped pexidartinib after C1D15 mainly due to toxicity. Histogram bars represent means +/- SEM. Data were analyzed using a student's *t*-test. **: $P < 0.01$.

**Supplementary Fig. 3: Pexidartinib does not impact the production of proinflammatory cytokines and the frequency and functions of T and B cells. (A)** Quantification by multiplex ECLIA of IL-1β, IL-6, and TNF-α in supernatants of patients' PBMCs after 6 hours stimulation with R848. Patients have either continuously taken pexidartinib (black symbols and bars) or have stopped pexidartinib after C1D15 mainly due to toxicity (grey symbols and bars). Histogram bars represent means +/- SEM. **(B)** Analysis of % $CD3^+$ T cells, CD4/CD8 ratio, and % $CD19^+$ B cells based on FC panels n°1 and 2 (**table S3**) are shown. Dots represent individual patients, whereas bars indicate median values. Red dotted lines represent healthy donors' median on each graph. Data were analyzed by a Wilcoxon test. **(C)** Quantification by multiplex ECLIA of IFN-γ and IL-2 in supernatants of patients' PBMCs after 6 hours stimulation with PMA and ionomycin. Dots represent individual patients, whereas bars indicate median values. Red dotted lines represent healthy donors' median on each graph. No statistical significance was reached using Student's *t*-test in (**A**) or a Kruskal-Wallis test followed by Dunn's correction in (**B-C**).

**Supplementary Fig. 4: Pexidartinib does not alter human DC viability**. PBMC from HD were treated for 16h with graded doses of pexidartinib (1µM-40µM) in complete RPMI medium (n=4). **(A)** Live cells, **(B)** Total CD11c$^+$ DC and **(C-E)** DC subsets (BDCA2$^+$ pDCs, BDCA1$^+$ cDC2, and BDCA3$^+$cDC1) were identified as previously described *(27)*. Histogram bars represent the fractions of (**A**) total live cells and (**B-E**) each population among live cells. For each panel, data represent mean ± SEM of 4 independent experiments and each symbol represents an individual experiment. No statistical significance was reached using one-way ANOVA test followed by Dunnett's correction.

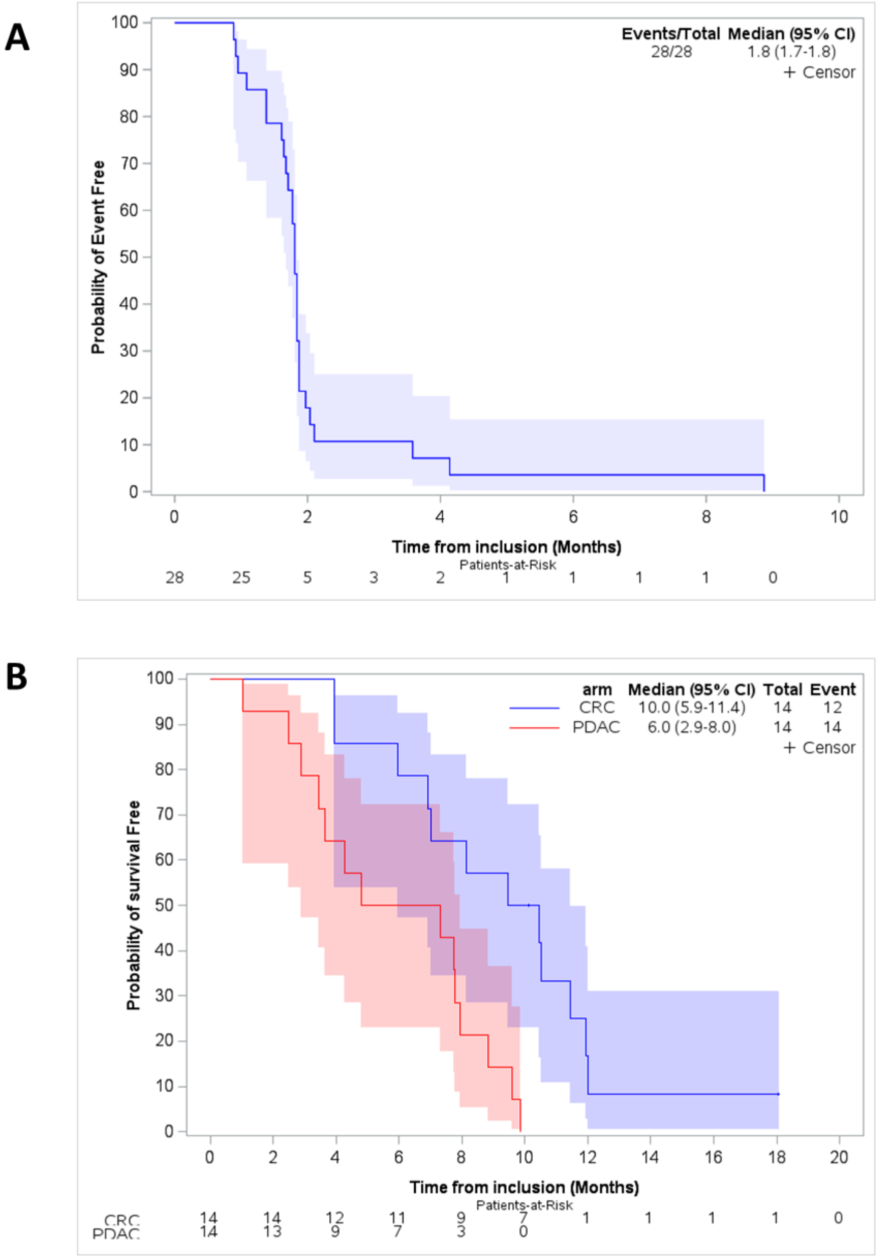


**Supplementary Fig. 1: Progression-Free Survival (PFS) and Overall Survival (OS).** Kaplan–Meier curves by RECIST version 1.1 for **(A)** PFS and **(B)** OS in 28 evaluable patients (expansion part) with advanced/metastatic CRC and PDAC. Median PFS and OS are indicated in months. Number of patients at risk at indicated time points are shown below the x-axis.

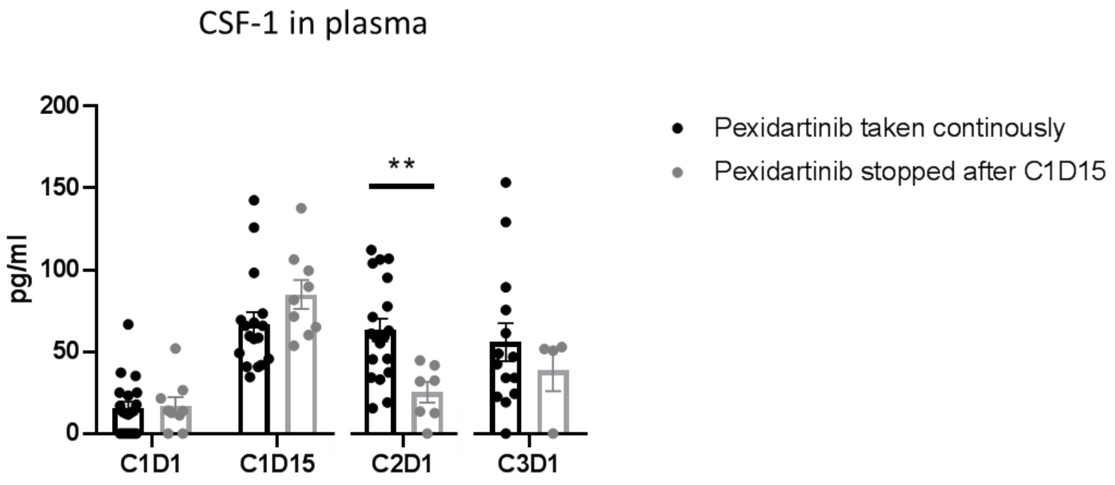


**Supplementary Fig. 2: Persistency of high CSF-1 levels only in patients who continued to receive pexidartinib.** CSF-1 levels were quantified by multiplex ECLIA in plasma of patients who have continuously taken pexidartinib versus those who have stopped pexidartinib after C1D15 mainly due to toxicity. Histogram bars represent means +/- SEM. Data were analyzed using a student's *t*-test. **: $P < 0.01$.

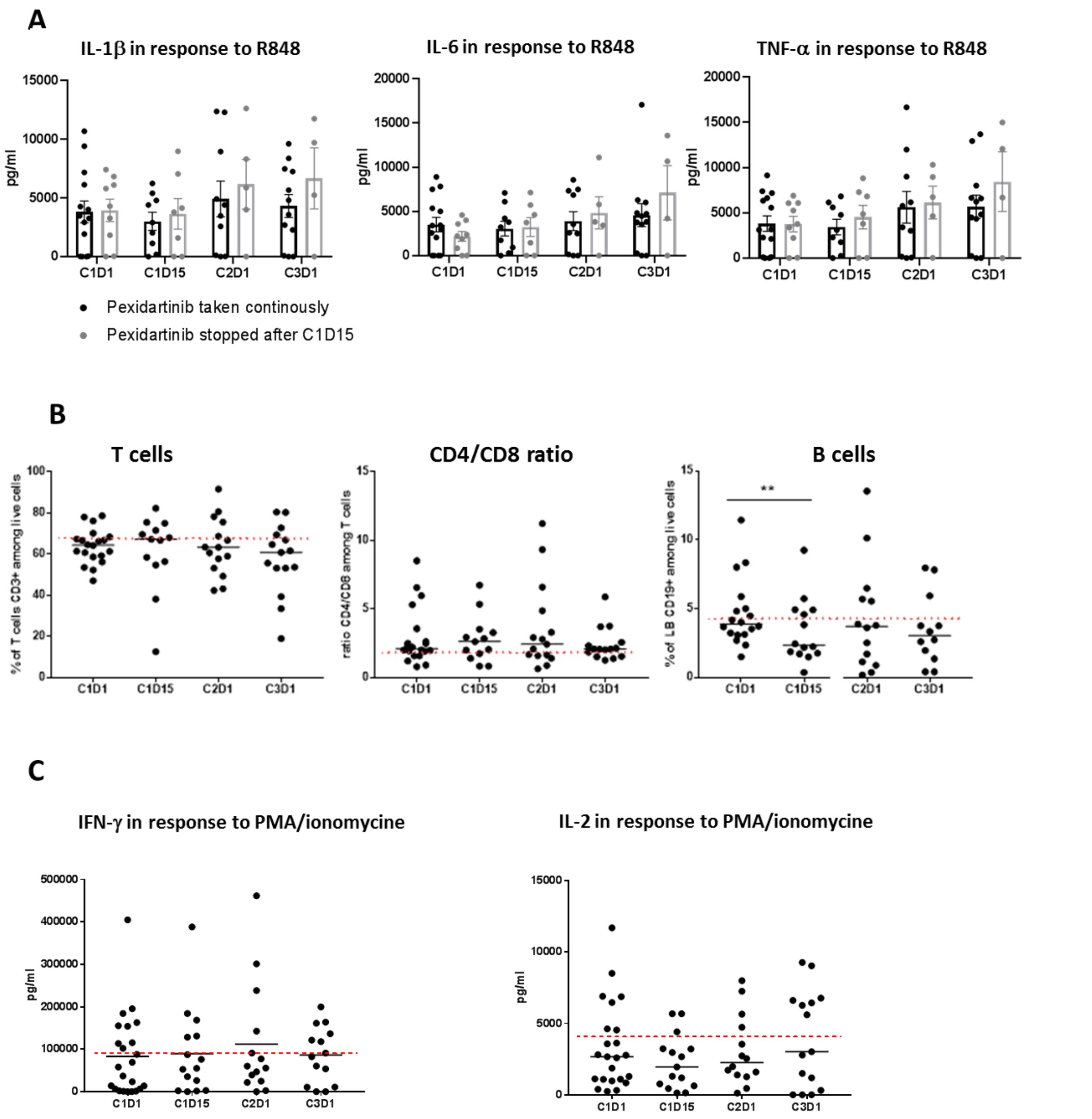


**Supplementary Fig. 3: Pexidartinib does not impact the production of proinflammatory cytokines and the frequency and functions of T and B cells. (A)** Quantification by multiplex ECLIA of IL-1β, IL-6, and TNF-α in supernatants of patients' PBMCs after 6 hours stimulation with R848. Patients have either continuously taken pexidartinib (black symbols and bars) or have stopped pexidartinib after C1D15 mainly due to toxicity (grey symbols and bars). Histogram bars represent means +/- SEM. **(B)** Analysis of % $CD3^+$ T cells, CD4/CD8 ratio, and % $CD19^+$ B cells based on FC panels n°1 and 2 (**table S3**) are shown. Dots represent individual patients, whereas bars indicate median values. Red dotted lines represent healthy donors' median on each graph. Data were analyzed by a Wilcoxon test. **(C)** Quantification by multiplex ECLIA of IFN-γ and IL-2 in supernatants of patients' PBMCs after 6 hours stimulation with PMA and ionomycin. Dots represent individual patients, whereas bars indicate median values. Red dotted lines represent healthy donors' median on each graph. No statistical significance was reached using Student's *t*-test in (**A**) or a Kruskal-Wallis test followed by Dunn's correction in (**B-C**).

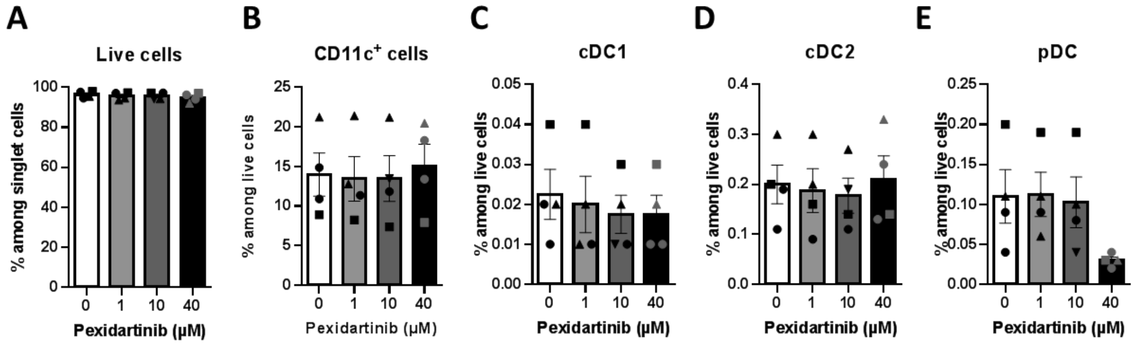


**Supplementary Fig. 4: Pexidartinib does not alter human DC viability**. PBMC from HD were treated for 16h with graded doses of pexidartinib (1µM-40µM) in complete RPMI medium (n=4). **(A)** Live cells, **(B)** Total CD11c$^+$ DC and **(C-E)** DC subsets (BDCA2$^+$ pDCs, BDCA1$^+$ cDC2, and BDCA3$^+$cDC1) were identified as previously described *(27)*. Histogram bars represent the fractions of (**A**) total live cells and (**B-E**) each population among live cells. For each panel, data represent mean ± SEM of 4 independent experiments and each symbol represents an individual experiment. No statistical significance was reached using one-way ANOVA test followed by Dunnett's correction.

**Supplementary table 1:** Treatment-Related Adverse Events (TRAE) observed during both parts. Data are n (%). Only events with frequency >5% are represented on the table. Adverse events and laboratory abnormalities were graded according to the National Cancer Institute Common Terminology Criteria for Adverse Events (NCI-CTCAE, version 4.03). Gr, Grade; AE, adverse event; ALT, alanine aminotransferase; AST, aspartate aminotransferase; ALP, alkaline phosphatase; ANC, absolute neutrophil count.

| | Dose escalation | | | | | | | | Expansion cohorts | | | | All | | | |
|---|---|---|---|---|---|---|---|---|---|---|---|---|---|---|---|---|
| | 400 mg N=3 | | 600 mg N=3 | | 800 mg N=3 | | 1000 mg N=10 | | N=28 | | | | N=47 | | | |
| | Gr 1-2 | Gr 3-4 | Gr 1-2 | Gr 3-4 | Gr 1-2 | Gr 3-4 | Gr 1-2 | Gr 3-4 | All grade | | Grade 3-4 | | All grade | | Grade 3-4 | |
| **Clinical AEs** | | | | | | | | | | | | | | | | |
| Fatigue | 2 | 1 | 1 | 0 | 1 | 0 | 6 | 2 | 22 | 79% | 5 | 18% | 35 | 74% | 8 | 17% |
| Peripheral edema | 0 | 0 | 3 | 0 | 5 | 0 | 6 | 0 | 9 | 32% | 0 | 0% | 23 | 49% | 0 | 0% |
| Anorexia | 1 | 0 | 0 | 0 | 0 | 0 | 5 | 0 | 16 | 57% | 1 | 4% | 22 | 47% | 1 | 2% |
| Diarrhea | 0 | 0 | 1 | 0 | 0 | 0 | 3 | 0 | 14 | 50% | 0 | 0% | 18 | 38% | 0 | 0% |
| Maculopaular rash | 0 | 0 | 1 | 0 | 2 | 0 | 5 | 0 | 8 | 29% | 4 | 14% | 16 | 34% | 4 | 9% |
| Nausea | 1 | 0 | 1 | 0 | 0 | 0 | 3 | 1 | 10 | 36% | 0 | 0% | 16 | 34% | 1 | 2% |
| Hair color changes | 1 | 0 | 1 | 0 | 0 | 0 | 6 | 0 | 4 | 14% | 0 | 0% | 12 | 26% | 0 | 0% |
| Pruritus | 0 | 0 | 0 | 0 | 1 | 0 | 2 | 0 | 9 | 32% | 0 | 0% | 12 | 26% | 0 | 0% |
| Vomiting | 0 | 0 | 0 | 0 | 0 | 0 | 4 | 0 | 8 | 29% | 0 | 0% | 12 | 26% | 0 | 0% |
| Dysgueusia | 0 | 0 | 1 | 0 | 0 | 0 | 1 | 0 | 4 | 14% | 0 | 0% | 6 | 13% | 0 | 0% |
| Dry skin | 0 | 0 | 1 | 0 | 2 | 0 | 0 | 0 | 3 | 11% | 0 | 0% | 6 | 13% | 0 | 0% |
| Dry mouth | 0 | 0 | 0 | 0 | 1 | 0 | 1 | 0 | 3 | 11% | 0 | 0% | 5 | 11% | 0 | 0% |
| Abdominal pain | 0 | 0 | 0 | 0 | 0 | 0 | 1 | 0 | 4 | 14% | 1 | 4% | 5 | 11% | 1 | 2% |
| Oral mucositis | 0 | 0 | 1 | 0 | 0 | 0 | 1 | 0 | 2 | 7% | 0 | 0% | 4 | 9% | 0 | 0% |
| Myalgia | 1 | 0 | 0 | 0 | 0 | 0 | 1 | 0 | 1 | 4% | 0 | 0% | 3 | 6% | 0 | 0% |
| **Investigations** | | | | | | | | | | | | | | | | |
| AST increase | 0 | 0 | 0 | 0 | 0 | 0 | 2 | 2 | 19 | 68% | 6 | 21% | 23 | 49% | 8 | 17% |
| ALT increase | 0 | 0 | 0 | 0 | 0 | 0 | 0 | 3 | 15 | 54% | 7 | 25% | 18 | 38% | 10 | 21% |
| Blood bilirubin increase | 1 | 0 | 0 | 0 | 0 | 0 | 1 | 1 | 4 | 14% | 0 | 0% | 7 | 15% | 1 | 2% |
| ALP increase | 0 | 0 | 0 | 0 | 0 | 0 | 0 | 3 | 3 | 11% | 3 | 11% | 6 | 13% | 6 | 13% |
| ANC decrease | 0 | 0 | 0 | 1 | 0 | 0 | 0 | 1 | 4 | 14% | 1 | 4% | 6 | 13% | 3 | 6% |
| Lymphocyte count decrease | 0 | 0 | 1 | 0 | 0 | 0 | 0 | 1 | 2 | 7% | 2 | 7% | 4 | 9% | 3 | 6% |
| Anemia | 0 | 0 | 0 | 0 | 0 | 0 | 1 | 0 | 2 | 7% | 0 | 0% | 3 | 6% | 0 | 0% |

**Supplementary table 2:** Treatment administration. Data are n (%). *: number of patient with at least one dose modification.

| | Dose Escalation | | | | Expansion Cohorts | | All | |
|---|---|---|---|---|---|---|---|---|
| | 400 mg N=3 | 600 mg N=3 | 800 mg N=3 | 1000 mg N=10 | N=28 | | N=47 | |
| **Pexidartinib** | | | | | | | | |
| Toxicity-related dose modification* | 0 | 1 | 1 | 6 | 21 | 75% | 29 | 62% |
| Permanent discontinuation | 3 | 3 | 3 | 10 | 28 | 100% | 47 | 100% |
| Reason of permanent discontinuation | | | | | | | | |
| Patient decision | 0 | 0 | 0 | 0 | 1 | 4% | 1 | 2% |
| Toxicity | 0 | 1 | 0 | 1 | 5 | 18% | 7 | 15% |
| Disease progression | 3 | 2 | 3 | 9 | 22 | 79% | 39 | 83% |
| **Durvalumab** | | | | | | | | |
| Permanent discontinuation | 3 | 3 | 3 | 9 | 28 | 100% | 46 | 98% |
| Reason of permanent discontinuation | | | | | | | | |
| Patient decision | 0 | 0 | 0 | 0 | 1 | 4% | 1 | 2% |
| Toxicity | 0 | 0 | 0 | 0 | 1 | 4% | 1 | 2% |
| Disease progression | 3 | 3 | 3 | 9 | 26 | 93% | 44 | 94% |

**Supplementary table 3:** Multiparametric FC panels used for the monitoring of the expansion cohort: **panel 1:** analysis of human B cells, DCs, and monocytes, **panel 2:** analysis of human T cells, **panel 3:** analysis of mouse DCs, and **panel 4**: analysis of human DC subsets.

| | **fluorochrome** | **antigen** |
|---|---|---|
| **panel 1** (human B cells, DCs, and monocytes) | BV480 | HLA-DR |
| | BV605 | CD19 |
| | BV650 | BDCA1 |
| | BV711 | CD14 |
| | PERCPVIO700 | CD16 |
| | PE | CD33 |
| | PEVIO615 | CD3 |
| | | CD56 |
| | PEVIO770 | CD11C |
| | APCVIO770 | BDCA2 |
| | | BDCA3 |
| | UV | Viability |
| **panel 2** (human T cells) | BV421 | KI67 |
| | BV480 | CD45RA |
| | BV650 | CD4 |
| | BV711 | CD8 |
| | FITC | CCR7 |
| | APC | FOXP3 |
| | AL700 | CD3 |
| | UV | Viability |
| **panel 3** (mouse DC) | BV711 | I-A/I-E |
| | BV650 | CD11b |
| | ZOMBIE aqua | Viability |
| | BV421 | XCR1 |
| | A488 | CD172 (SIRPa) |
| | PE Cy7 | CD11c |
| | PE-Dazzle 594 | CD64 |
| | PE | BST2 |
| | APC-Cy7 | Ly6c |
| | APC | SiglecH |
| **panel 4** (human DC within PBMCs) | BV480 | HLA-DR |
| | BV650 | CD1C |
| | BV711 | CD14 |
| | PERCPVIO700 | CD16 |
| | PEVIO770 | CD11c |
| | APC | BDCA2 |
| | | BDCA3 |
| | AL700 | CD3 |
| | | CD15 |
| | | CD19 |
| | LD ZOMBIE Near-IR | Viability |

**Supplementary table 4:** Multiparametric immunofluorescence stainings using specific surface marker combinations.

| | Primary antibodies | | | | | Secondary antibodies - Fluorophore | | | |
|---|---|---|---|---|---|---|---|---|---|
| **Target** | **Species - Isotype** | **Provider** | **Reference** | **Clone** | **Dilution** | **Host species and conjugate** | **Reference** | **Provider** | **Dilution** |
| **CD68** | Mouse IgG2b | R&D | MAB20401 | 298807 | 1/50 | Goat anti-Mouse IgG2b AF647 | A21242 | Invitrogen | 1/500 |
| **CD163** | Mouse IgG1 | Leica | NCL-L-CD163 | 10D6 | 1/30 | Goat anti-Mouse IgG1 AF488 | A21121 | Invitrogen | |
| **CK** | Mouse IgG1 | DAKO | M3515 | AE1/AE3 | 1/50 | Goat-anti Mouse IgG AF488 | A11029 | Invitrogen | 1/500 |
| **MPO** | Rabbit IgG | DAKO | A0398 | polyclonal | 1/100 | Goat-anti Rabbit IgG AF647 | A21245 | Invitrogen | |
| **CD3** | Rabbit IgG | DAKO | A0452 | polyclonal | 1/300 | Goat Anti-Rabbit IgG AF647 | A21245 | Invitrogen | 1/500 |
| **CD20** | Mouse IgG2a | DAKO | M0755 | L26 | 1/400 | Goat Anti-Mouse IgG AF555 | A21424 | Invitrogen | |
| **CD8** | Mouse IgG1 | DAKO | M7103 | C8/144B | 1/30 | Goat-anti Mouse IgG AF647 | A21236 | Invitrogen | 1/500 |
| **Ki67** | Rat IgG | e-Biosciences | 14-5698-80 | SolA15 | 1/70 | Goat anti-Rat IgG AF555 | A21434 | Invitrogen | |
| **CD4** | Rabbit IgG | Ventana | 790-4423 | polyclonal | Ready-to-use | Goat-anti Rabbit IgG AF488 | A11034 | Invitrogen | |
